# A comparison of ten polygenic score methods for psychiatric disorders applied across multiple cohorts

**DOI:** 10.1101/2020.09.10.20192310

**Authors:** Guiyan Ni, Jian Zeng, Joana A Revez, Ying Wang, Zhili Zheng, Tian Ge, Restuadi Restuadi, Jacqueline Kiewa, Dale R Nyholt, Jonathan R I Coleman, Jordan W Smoller, Schizophrenia Working Group of the Psychiatric Genomics Consortium, Major Depressive Disorder Working Group of the Psychiatric Genomics Consortium, Jian Yang, Peter M Visscher, Naomi R Wray

## Abstract

**Background:** Polygenic scores (PGSs), which assess the genetic risk of individuals for a disease, are calculated as a weighted count of risk alleles identified in genome-wide association studies (GWASs). PGS methods differ in which DNA variants are included and the weights assigned to them; some require an independent tuning sample to help inform these choices. PGSs are evaluated in independent target cohorts with known disease status. Variability between target cohorts is observed in applications to real data sets, which could reflect a number of factors, e.g., phenotype definition or technical factors.

**Methods:** The Psychiatric Genomics Consortium working groups for schizophrenia (SCZ) and major depressive disorder (MDD) bring together many independently collected case-control cohorts. We used these resources (31K SCZ cases, 41K controls; 248K MDD cases, 563K controls) in repeated application of leave-one-cohort-out meta-analyses, each used to calculate and evaluate PGS in the left-out (target) cohort. Ten PGS methods (the baseline PC+T method and nine methods that model genetic architecture more formally: SBLUP, LDpred2-Inf, LDpred-funct, LDpred2, Lassosum, PRS-CS, PRS-CS-auto, SBayesR, MegaPRS) are compared.

**Results:** Compared to PC+T, the other nine methods give higher prediction statistics, MegaPRS, LDPred2 and SBayesR significantly so, up to 9.2% variance in liability for SCZ across 30 target cohorts, an increase of 44%. For MDD across 26 target cohorts these statistics were 3.5% and 59%, respectively.

**Conclusions:** Although the methods that more formally model genetic architecture have similar performance, MegaPRS, LDpred2, and SBayesR rank highest in most comparison and are recommended in applications to psychiatric disorders.

## Introduction

Polygenic scores (PGSs), which assess the genetic risk of individuals for a disease(1, 2), are calculated as a weighted count of genetic risk alleles in the genome of an individual, with the risk alleles and their weights derived from the results of genome-wide association studies (GWAS)(3). PGS can be calculated for any trait or disease with sufficiently powered GWAS (‘discovery samples’), and accuracy of PGS applied in independent GWAS ‘target samples’ will increase as discovery sample size increases. Since genetic factors only capture the genetic contribution to risk and since PGS only capture part of the genetic risk, PGS cannot be diagnostically accurate risk predictors (see review(4)). Nonetheless, for many common complex genetic disorders, such as cancers(5, 6) and heart disease(7, 8), there is increasing interest in evaluating PGS for early disease detection, prevention and intervention(9–11).

There are now many methods to calculate PGSs, and the methods differ in terms of two key criteria: which DNA variants to include and what weights to allocate to them. Here, for simplicity, we assume the DNA variants are single nucleotide polymorphisms, SNPs, but other DNA variants tested for association with a trait can be used. While stringent thresholds are set to declare significance for association of individual SNPs in GWAS, PGSs are robust to inclusion of some false positives. Hence, the maximum prediction from PGSs tested in target samples may include nominally associated SNPs. The optimum method to decide which SNPs to select and what weights to allocate them, may differ between traits depending on the sample size of the discovery GWAS and on the genetic architecture of the trait (the number, frequencies and effect sizes of causal variants), particularly given the linkage disequilibrium (LD) correlation structure between SNPs. Often, when new PGS methods are introduced, comparisons are made between a limited set of methods using simulated data, together with application to some real data examples. However, it can be difficult to compare across the new methods, particularly because in real data there can be variability in PGS evaluation statistics between target cohorts, not encountered in idealised simulations. The reasons for this variability are usually unknown and not simple to identify (12) but could reflect a number of factors such as phenotype definition, ascertainment strategies of cases and controls, cohort-specific ancestry within the broad classification of ancestry defined by the GWAS discovery samples (e.g., European), or technical artefacts in genotype generation.

Here, we compare ten PGS methods (PC+T(3, 13), SBLUP(14), LDpred2-Inf(15), LDpred2(15), LDpred-funct(16), Lassosum(17), PRS-CS(18), PRS-CS-auto(18) and SBayesR(19), MegaPRS(20), **Table 1**). Some of these methods (PC+T, LDpred2, MegaPRS, Lassosum and PRS-CS) require a ‘tuning sample’, a GWAS cohort with known trait status that is independent of both discovery and target samples, used to select parameters needed to generate the PGSs in the target sample. Whereas only GWAS summary statistics are needed for discovery samples, individual level genotype data are needed for tuning and target samples. Information about the LD structure is supplied by a reference data set of genome-wide genotypes which can be independently collected from the GWAS data, but from samples of matched ancestry.

**Table 1.**
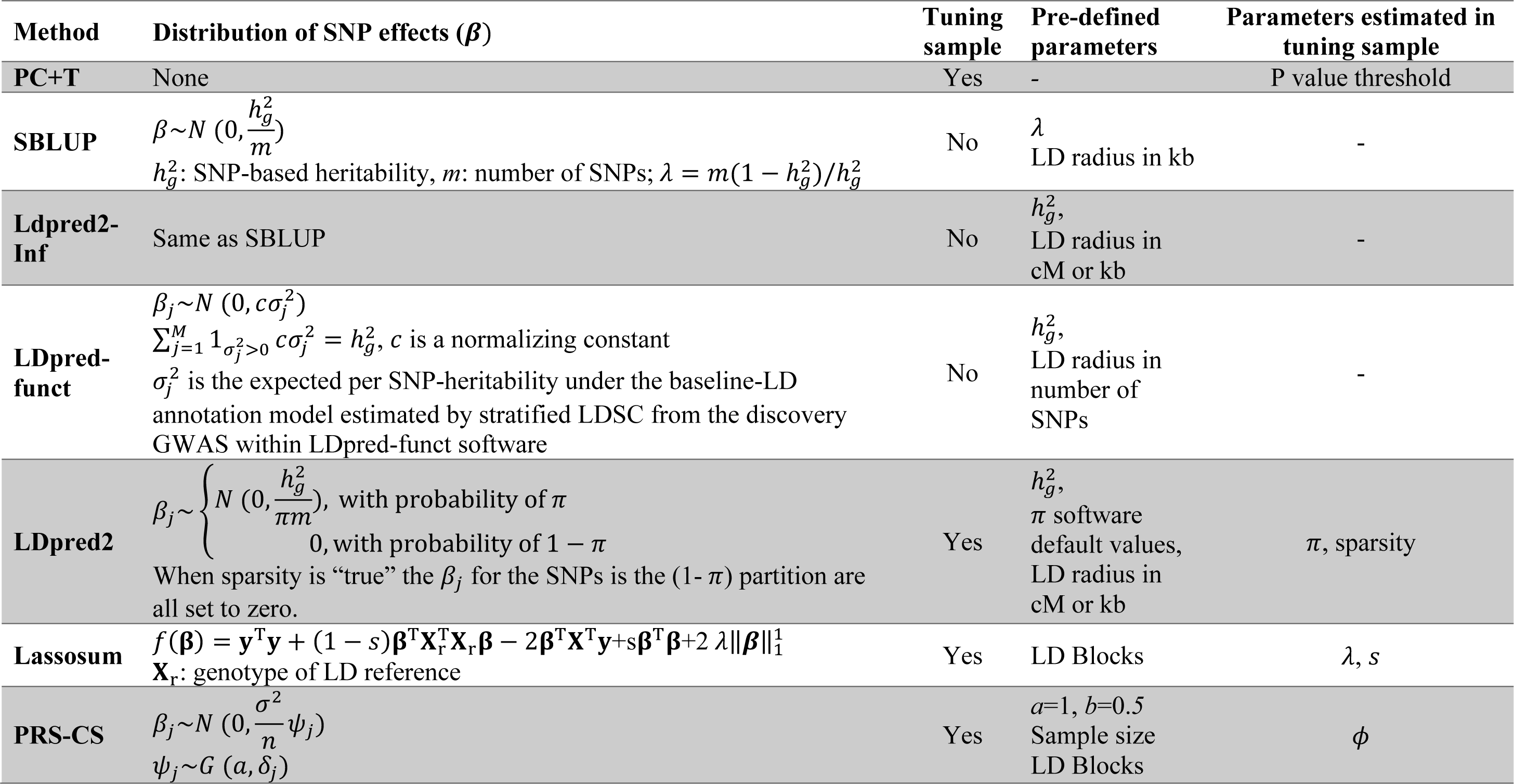
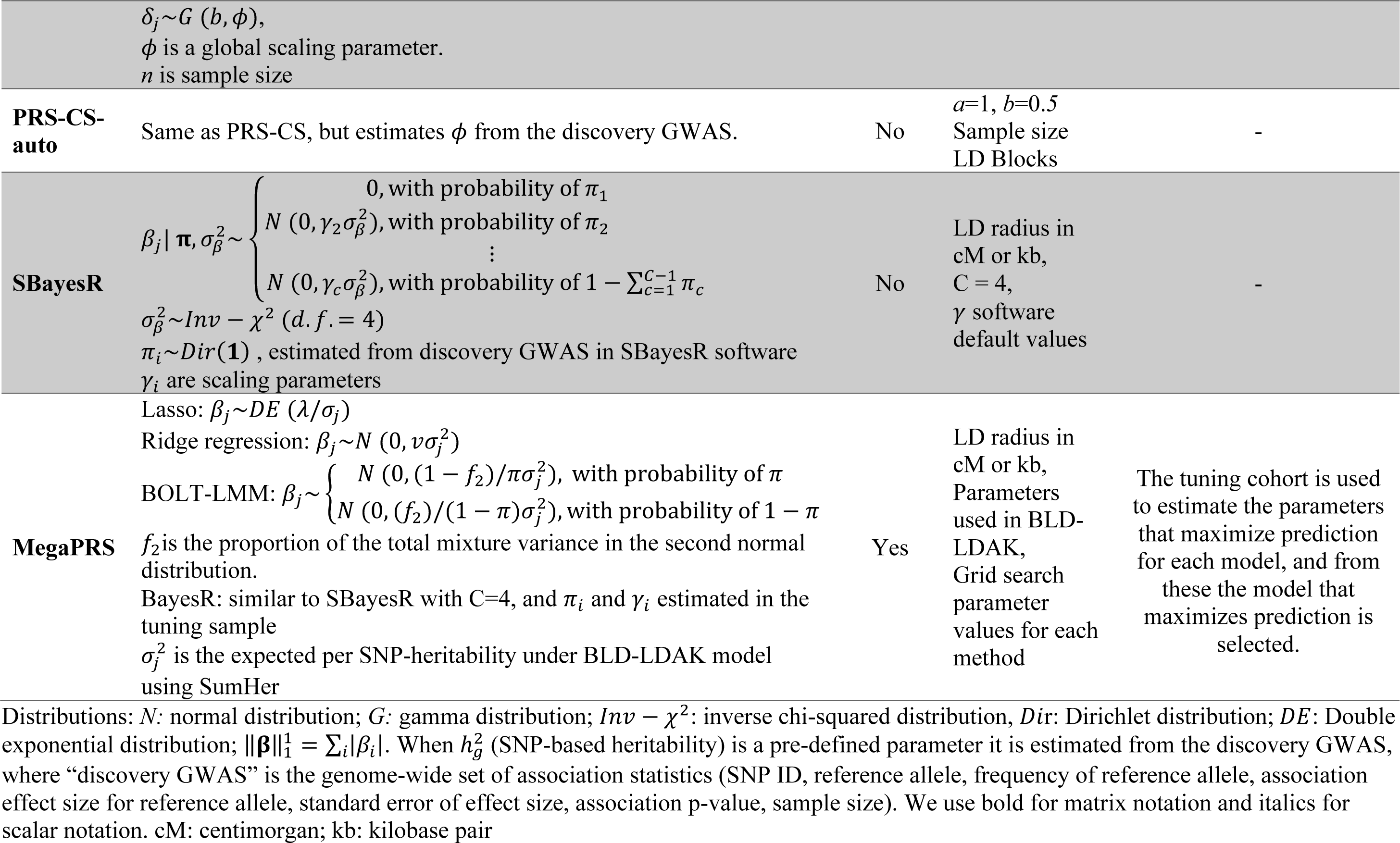
Summary of methods used to generate PGS

Briefly, PC+T (P-value based clumping and thresholding, also known as the P+T or C+T method) uses the GWAS effect size estimates as SNP weights and includes independent SNPs (defined by an LD *r*^2^ filter for a given chromosomal window distance) with association P-values lower than a threshold (chosen after application in a tuning sample). PC+T is the most commonly used and basic method, and so is the benchmark method here. The other methods assume either that all SNPs have an effect size drawn from a normal distribution (SBLUP and LDpred2-Inf) or that SNP effects are drawn from mixtures of distributions with the key parameters defining these architectures estimated through Bayesian frameworks (LDpred2, PRS-CS, SBayesR). LDpred-funct and MegaPRS include functional annotation to SNPs to up/down weight their contributions to the PGSs, which could improve prediction accuracy if this functional information helps to better separate true and false positive associations(21). The MegaPRS software implements a suite of methods (**Table 1**) and selects the method, together with its parameter estimates, that maximises prediction in the tuning cohort. MegaPRS utilises the BLD-LDAK model(22) where the variance explained by each SNP depends on its allele frequency, LD and functional annotations. Notably, some methods (SBayesR, PRS-CS-auto and LDpred2-auto) do not require a tuning cohort, so that the SNPs selected and their weights reflect only the properties of the discovery sample. Since LDpred2-auto is shown to perform similarly to LDpred2, we do not include it in comparisons made here. We apply these methods to data from the Psychiatric Genomics Consortium (PGC) working groups for schizophrenia (SCZ)(23, 24) and major depressive disorder (MDD)(12, 25, 26) (**Tables S1 and S2**). We select SCZ and MDD to study as they have the largest GWAS samples for psychiatric disorders to date but are diverse in lifetime risk, and are representative of psychiatric disorders which have all been shown to be highly polygenic (27). The PGC provides a useful resource for undertaking this study because it brings together many independently collected cohorts for GWAS meta-analysis. This allows the application of repeated leave-one-cohort-out GWAS analyses generating robust conclusions from evaluation of PGS applied across multiple left-out target cohorts.

## Materials and Methods

### Data

All samples were of European ancestry with full details in the **Supplementary Note, Table S1** and **S2**. Briefly, GWAS summary statistics were available from PGC SCZ for 37 European ancestry cohorts (24) (31K SCZ cases and 41K controls) of which 34 had individual level data available. PGS were calculated in each of the 30 cohorts (target samples) using the GWAS discovery sample based on a meta-analysis of 37-2 = 35 cohorts (24) i.e., the target sample was excluded from the discovery sample as well as a sample selected to be a tuning sample. Analyses were repeated using four different tuning samples, two of which were large (swe6:2313; gras: 2318) and two were small (lie2:406; msaf:466). Similarly, GWAS MDD summary statistics were available from 248K cases and 563K controls(25), which included data from the 26 cohorts from PGC MDD with individual level data (15K cases and 24K controls). We left one cohort out of those 26 cohorts in turn as the target sample, and then used a meta-analysis of remaining data as discovery samples. A cohort(25), not included in the discovery GWAS was used as the tuning sample (N=1,679).

### Baseline SNP selection

For baseline analyses, only SNPs with minor allele frequency (MAF) > 0.1 and imputation INFO score > 0.9 (converted to best-guess genotype values of 0, 1 or 2) were selected.

Sensitivity analyses relaxed the MAF threshold to MAF > 0.05 or 0.01 and INFO score threshold to 0.3. All methods were conducted using HapMap3 SNPs, except the method PC+T, which was conducted based on all imputed SNPs (8M in SCZ, and 13M in MDD).

### Prediction methods

We define a PGS of an individual, *j*, as a weighted sum of SNP allele counts: 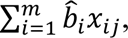, where *m* is the number of SNPs included in the predictor, *m* is the per allele weight for the SNP, *x_ij_*, is a count of the number (0, 1, or 2) of trait-associated alleles of SNP *i* in individual *j*. We compared ten risk prediction methods, described in the **Supplemental Note** and summarized in **Table 1**. The methods differ in terms of the SNPs selected for inclusion in the predictor and the *b*^*_i_* values assigned to the SNPs. All methods use the GWAS summary statistics as the starting point, but each makes choices differently for which SNPs to include and for the *b*^*_i_* values to assign. Some methods use a tuning cohort; parameter estimates that maximize prediction in that tuning cohort are selected for application in the target sample. Several methods employ an LD reference sample to infer the expected correlation structure between SNP association statistics, those recommended by each software implementation are used.

### Evaluation of out-of-sample prediction

The accuracy of prediction in each target cohort was quantified by 1) Area under the receiver operator characteristic curve (AUC; R library pROC(35)). AUC can be interpreted as a probability that a case ranks higher than a control. 2) The proportion of variance on the liability scale explained by PGS(36). We used the population lifetime risk of SCZ and MDD as 1% and 15% respectively to convert the variance explained in a linear regression to the liability scale(25, 28, 37). 3) Odds ratio (OR) of tenth PGS decile relative to the first decile. 4) Odds ratio of tenth PGS decile relative to those ranked in the middle of the PGS distribution, which is calculated as the average of OR of tenth decile relative to fifth and sixth decile. 5) Standard deviation unit increase in cases. The PGS in each target cohort were scaled by standardising the PGS of controls and applying the standardisation to cases: 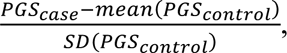, where SD is standard deviation. This does not impact PGS evaluation statistics but simply means that PGS are in SD units for all cohorts. The regression analyses for evaluation statistics 2-4 include 6 ancestry principal components as covariates.

These covariates are not included in the AUC model and the standard deviation unit increase in cases model (see **Supplementary Note**).

## Results

Prediction evaluation statistics based on all ten PGS methods and applied to SCZ across 30 study cohorts (**Figure 1, Figure S1, Table S3** and **S4**), and to MDD across 26 cohorts (**Figure S2, Table S5** and **S6**) are presented. There is variability in prediction statistics across target cohorts (as observed before(12, 28)) which is not a reflection of sample size (**Figure S3** and **Table S4** for SCZ, **Figure S4** and **Table S6** for MDD). Some significant associations were found from regression of prediction statistics on principal components (PCs) estimated from genome-wide SNPs (for SCZ **Figure S3**, but not MDD **Figure S4**), where the PCs capture both within-European ancestry and array differences between cohorts. The correlations of PGS between different methods are high (**Table S7**), but are lowest between PC+T and other methods (minimum 0.68). In contrast, the correlations between the other nine methods are always > 0.82. In theory, LDpred2-Inf and SBLUP are the same method. In practice, there are differences in implementation (e.g., different input parameters associated with definition of LD window) and although the correlation between their PGS is 0.974 the prediction accuracy is consistently higher for LDpred2-Inf. For SCZ, the AUC for all nine methods that directly model genetic architecture, other than PRS-CS-auto, are significantly higher than the PC+T method at the nominal level (**Figure 1A**). PGS from LDpred2, SBayesR and MegaPRS are significantly higher than the PC+T method after Bonferroni correction (p-value < 0.0011=0.05/45 (45 pairwise comparisons between 10 methods), one-tailed Student’s t-test). For MDD none of the differences between methods were significant (**Figure S2A**). For both SCZ and MDD across all statistics, regardless of tuning cohorts, LDpred2, SBayesR and MegaPRS, show relatively better performance (median across target cohorts) than other methods, although there is no significant difference between the nine methods that directly model genetic architecture. For variance explained on the liability scale, the PC+T PGS explained 6.4% for SCZ, averaged over the median values across the four tuning cohorts (**Figure 1B**), while it was 8.9%, 9.0%, and 9.2% for MegaPRS, LDpred2, and SBayesR, corresponding to an increase of 39%, 41% and 44%, respectively. For MDD although the variance explained is lower in absolute terms, 2.2% for PC+T *vs* 3.4% for MegaPRS, 3.5% for LDpred2 and 3.5% for SBayesR; the latter represents a 59% increase (**Figure S2B**).

**Figure 1.**
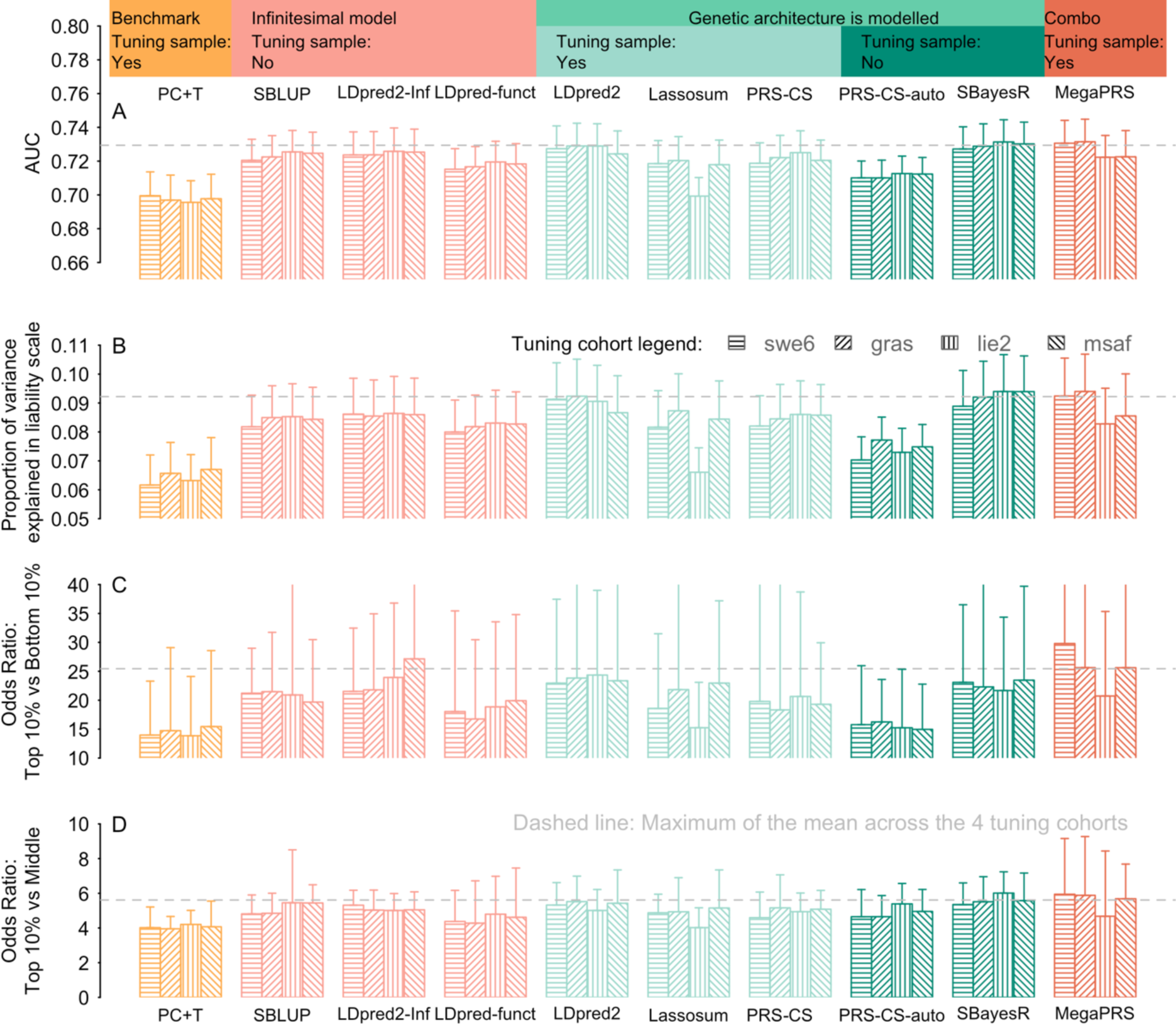
Prediction results for SCZ case/control status using different PGS methods. The PGS were constructed from SCZ GWAS summary statistics excluding the target cohort and a tuning cohort (shading legend). Each bar reflects the median across 30 target cohorts, the whiskers show the 95% confidence interval for comparing medians. The area under curve (AUC) statistic (A) can be interpreted as the probability that a case ranks higher than a control. Panel (B) is the proportion of variance explained by PGS on the scale of liability, assuming a population lifetime risk of 1%. The third panel (C) is the odds ratio when considering the odds of being a case comparing the top 10% vs bottom 10% of PGS. The bottom panel (D) is the odds of being a case in the top 10% of PGS vs odds of being a case in the middle of the PGS distribution. The middle was calculated as the averaged odds ratio of the top 10% ranked on PGS relative to the 5th decile and 6th decile. PC+T (also known as P+T) is the benchmark method which is shown in orange. Pink shows the methods that use an infinitesimal model assumption. The green shows the methods that model the genetic architecture, with light green for the methods using a tuning cohort to determine the genetic architecture of a trait; dark green shows the methods learning the genetic architecture from discovery sample, without using a tuning cohort. Dark orange is for MegaPRS using the BLD-LDAK model that assume the distribution of SNP effect depends on its allele frequency, LD and function annotation. MegaPRS assign four priors to each of SNP: LASSO, Bridge, BOLT-LMM, BayesR. Each prior has different hyperparameters that identified using the tuning cohort. The dashed grey lines are the maximum of the average across the four tuning cohorts. The sample sizes of the tuning cohorts are swe6: 1094 cases,1219 controls; lie2: 137 cases, 269 controls; msaf: 327 cases, 139 controls; gras: 1086 cases, 1232 controls.

We provide several evaluation statistics that focus on those in the top 10% of PGS, because clinical utility of PGS for psychiatric disorders is likely to focus on individuals that are in the top tail of the distribution of predicted genetic risk. The odds ratio for top *vs* bottom decile are large, ranging from 14 for PC+T to 30 for MegaPRS for SCZ and 3 for PC+T to 3.7 for SBayesR for MDD. While these top *vs* bottom decile odds ratios (**Figure 1C** and **S2C**) are much larger than the odds ratio obtained by using PGS to screen a general population (**Figure 1D** and **2D**) or patients in a healthcare system to identify people at high risk(38, 39), these comparisons are useful for research purposes, which could, for example, make cost-effective experimental designs focussing on individuals with high *vs* low PGS(40). The odds ratio of top 10% *vs* middle 10% are much less impressive, up to median of 6 for SCZ and 2 for MDD, but more fairly represents the value of PGS in population settings. These values can be benchmarked against risk in 1^st^ degree relatives of those affected, which are of the order of 8 for SCZ and 2 for MDD; low values are always expected for MDD because it is more common (lifetime risk ∼15% compared to ∼1% for SCZ). The odds ratio values are particularly high for some cohorts (**Table S4**), because in some SCZ cohorts the bottom 10% include very few or no cases, especially in cohorts with relatively small sample sizes.

**Figure 2.**
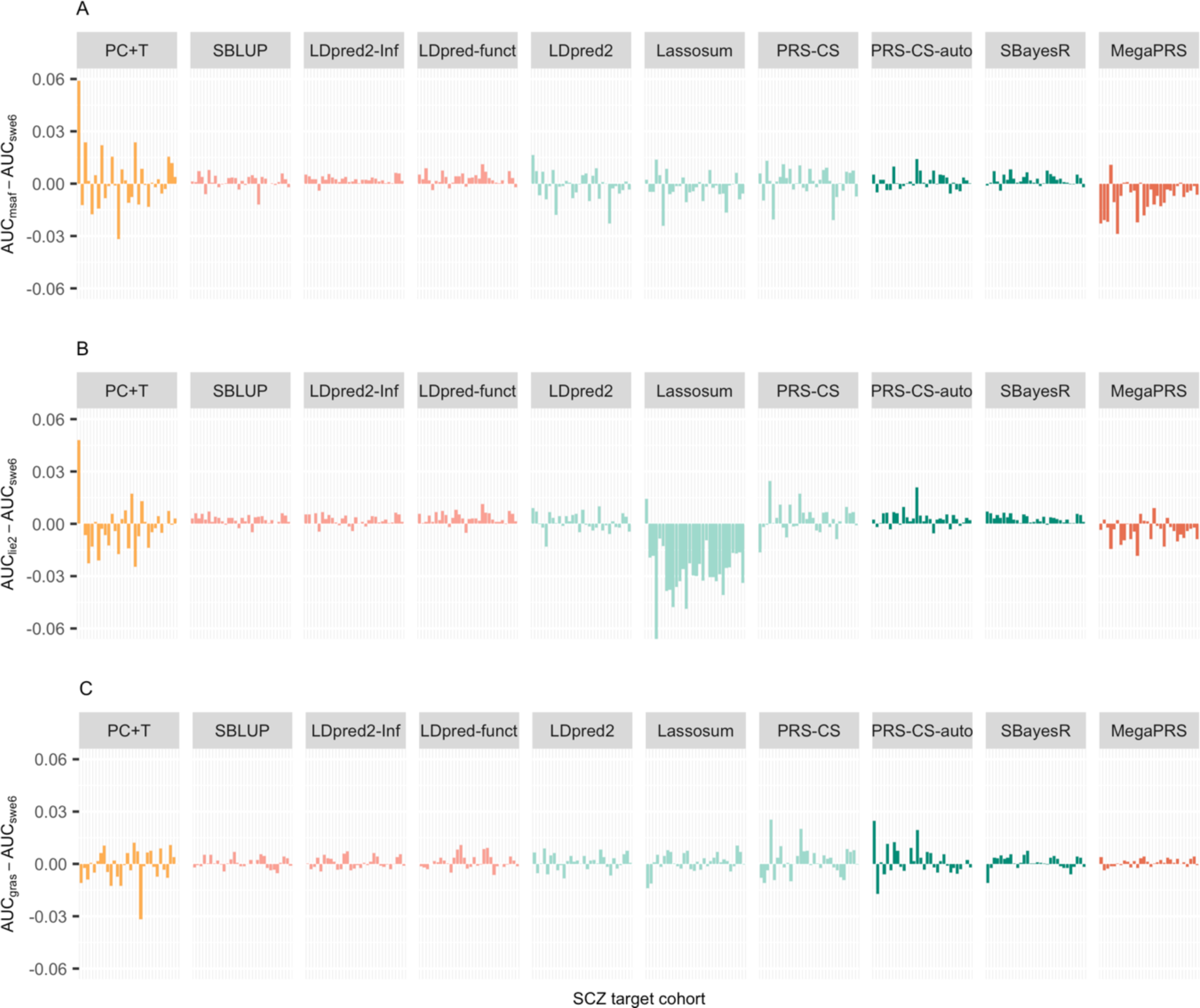
Sensitivity analyses using different tuning cohorts comparing different PGS methods. Differences in the AUC of SCZ of a PGS method when using different tuning cohorts. The different bars in each method (x-axis) refer to different validation cohorts ordered by sample size. The y-axis is the AUC difference when using alternative tuning cohort (i.e. lie2 (137 cases, 269 controls), msaf (327 cases, 139 controls), or gras (1086 cases, 1232 controls)), compared to ‘swe6’ (1094 cases, 1219 controls). The MAF QC threshold is 0.1. Note: SBLUP, LDpred2-Inf and LDpred-funct, PRS-CS-auto and SBayesR do not need a tuning cohort, but serve as a benchmark to the other methods which need a tuning cohort. These methods differ when a different tuning cohort is left out because the discovery GWAS also changes.

### The impact of tuning cohort

Five methods (i.e., PC+T, LDpred2, Lassosum, PRC-CS, and MegaPRS) use tuning cohorts to determine key parameters for application of the method into the target cohorts. Tuning parameters impact results in two ways. First, the parameters may be dependent on the choice of tuning cohort. Second, the discovery GWAS sample may be reduced in size (and hence power) if a tuning cohort needs to be excluded from the discovery GWAS. In all our analyses the tuning cohort is excluded from all GWAS discovery samples so that GWAS discovery sample is not variable across methods for each target cohort. Our results show that the tuning cohort can have considerable impact (**Figures 1, 2**). In our results, the tuning cohort that generates higher PGS is method dependent and differs between cohorts. For the methods that use tuning samples, the larger tuning samples (swe6 and gras) mostly generate higher prediction statistics compared to the two smaller tuning samples (lie and msaf), but the differences are not statistically significant. Although methods SBLUP, LDpred2-Inf, LDpred-funct, PRS-CS-auto and SBayesR require no tuning cohort, they serve as a benchmark, since the differences in their results reflect differences in the changed discovery samples (e.g., msaf is in the discovery sample, when swe6 is the tuning cohort, and *vice versa*), as well as the stochasticity inherent in the Gibbs sampling of Bayesian methods.

### The impact of MAF/INFO threshold

A MAF threshold of 0.1 and a INFO threshold of 0.9 are used to be consistent with applications in the PGC SCZ(28) and PGC MDD(25) studies, which had been imposed recognising that these thresholds generated more robust PGS results than using lower threshold values. In the second sensitivity analysis applied to the SCZ data, the MAF threshold was relaxed to 0.05 or 0.01 (**Figure 3**). The prediction evaluation statistics increase for some cohorts and decrease for others (trends with sample size were not significant). PC+T is more impacted that the other nine methods. Across target cohorts, different evaluation statistics were almost identical when including less common SNPs (**Table S3**). Relaxing the INFO score to 0.3 has a negligible effect (**Figure S5**).

**Figure 3.**
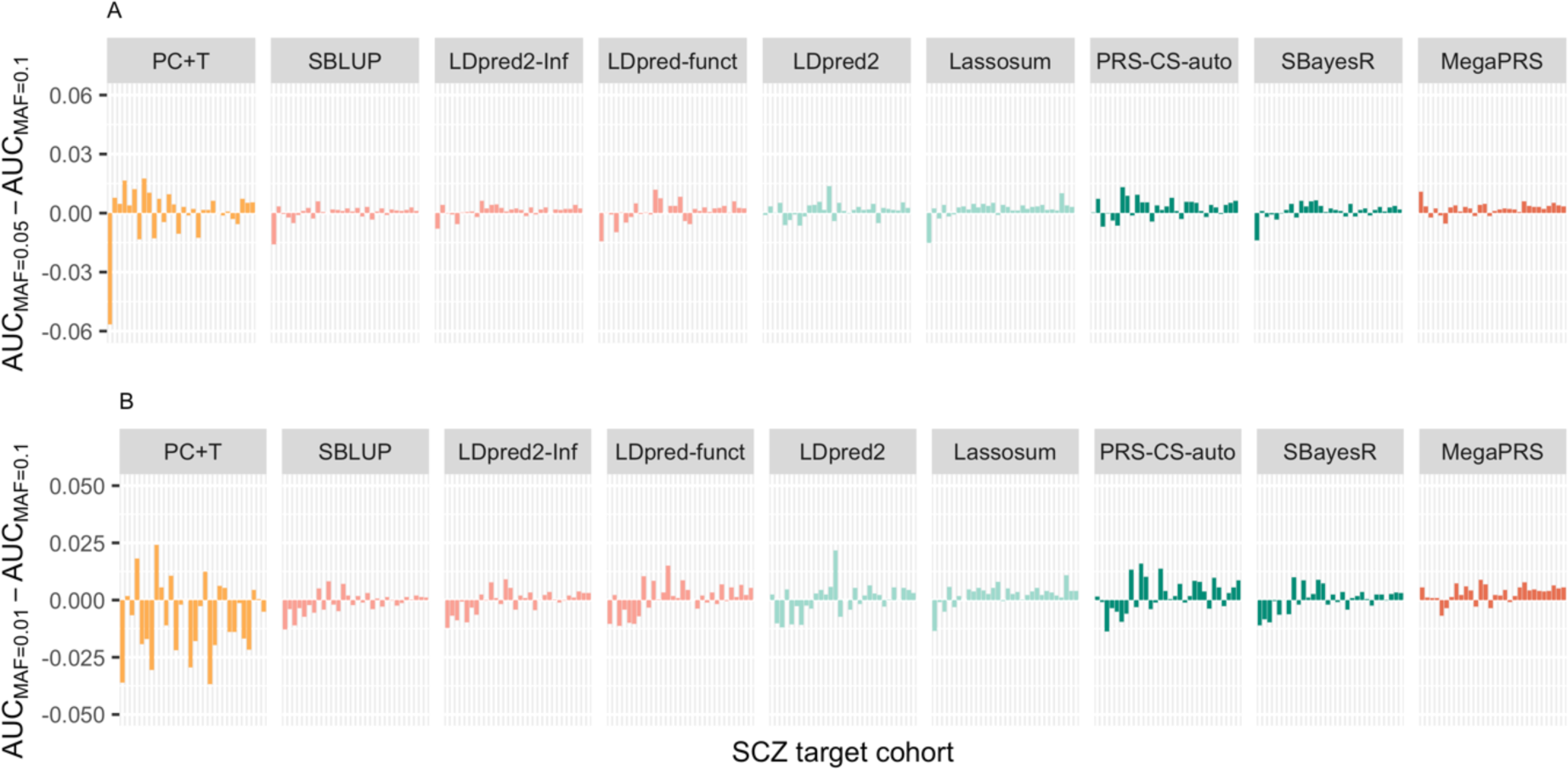
Sensitivity analyses using different MAF quality control thresholds. Differences in AUC of SCZ of a PGS method when using different MAF QC thresholds. The different bars in each method (x-axis) refer to different validation cohorts ordered by sample size. The y-axis is the AUC difference between analyses using A) MAF<0.05 and MAF <0.1 B) MAF<0.01 and MAF <0.1 as a QC threshold. The tuning cohort is ‘swe6’.

## Discussion

Comparison of PGS risk prediction methods showed that all nine methods that directly model genetic architecture had higher prediction evaluation statistics over the benchmark PC+T method for SCZ and MDD. While the differences between these nine methods were small, we found that MegaPRS, LDpred2, and SBayesR consistently ranked highest. Given that the PGS is a sum of many small effects, a normal distribution of PGS in a population is expected (and observed **Figures S6-9**). In idealised data, such as the relatively simple simulation scenarios usually considered in method development, all evaluation statistics should rank the same, but with real data sets this is not guaranteed. This is the motivation for considering a range of evaluation statistics. Our focus on statistics for those in the top 10% of PGS is relevant to potential clinical utility. In the context of psychiatry, it is likely that this will focus on people presenting in a prodromal state with clinical symptoms that have not yet specific to a diagnosis(11, 41). High PGS in those presenting to clinics could help contribute to clinical decision-making identifying individuals for closer monitoring or earlier intervention. Since a genetic-based predictor only predicts part of the risk of disease, and since a PGS only predicts part of the genetic contribution to disease it is acknowledged that PGS cannot be fully accurate predictors. Hence, the discriminative ability of PGS is low in the general population and the use of PGS in clinical settings requires evaluation including related ethical issues (42). Nonetheless, PGS, in combination with clinical risk factors, could make a useful contribution to risk prediction(41, 43, 44).

In sensitivity analyses that used different quality criteria for SNPs, e.g. MAF of 0.01 *vs* 0.05, INFO of 0.3 *vs* 0.9, we concluded that, currently, there is little to be gained in PGS from including SNPs with MAF < 0.10 and INFO < 0.9 for the diseases/dataset studied (**Table S8** and **S9**). This result may seem counter-intuitive since variants with low MAF are expected to play an important role in common disease, and some may be expected to have larger effect sizes than more common variants(45, 46). However, sampling variance is a function of allele frequency (≈ var (*y*)/ (2*MAF (1-MAF)\**n*)), where *y* is the phenotype and *n* is sample size), such that a variant of MAF =0.01 has sampling variance 9 times greater than a variant of MAF=0.1. Moreover, in real data sets small sample size of contributing cohorts mean that technical artefacts can accumulate to increase error in effect size estimates particularly of low frequency variants. Our conclusion that little is gained from including variants of MAF < 0.1 and reducing INFO threshold needs to be revisited as larger individual cohorts in discovery samples and larger target cohorts accumulate. Moreover, our comparison of methods uses only study samples of European ancestry. More research and data are needed to understand the properties of prediction methods within other ancestries and across ancestries, given potential differences in genetic architectures (in terms of number, frequencies and effect sizes of causal variants) and LD between measured variants and causal variants(47, 48).

For both SCZ and MDD, while the methods other than PC+T had similar performance, LDpred2, MegaPRS, and SBayesR saw the highest prediction accuracy in most of the comparisons. We note that we did not consider a version of PC+T that has been shown to have higher out of sample prediction compared to the standard implementation(13). This method conducts a grid search in a tuning cohort to determine LD *r*^2^ and INFO score thresholds for SNPs as well as the P-value threshold. Since the optimum LD threshold is likely to vary across genomic regions, the grid search approach is less appealing than the methods which implicitly allow this to vary. A sensitivity analysis in which we varied the *r*^2^ threshold in the PC+T showed only a small gain from optimising this (**Table S10**). LDpred2 has a version that does not require a tuning sample, LDpred2-auto, but the authors showed the two methods give similar results. SBayesR assumes that the SNP effects are drawn from a mixture of four distributions, which allows more flexibility in distributions of SNP effects by varying the proportion of SNPs in each distribution. Hence, SBayesR can fit essentially any underlying architecture in term of variance explained by each SNP so that the SBLUP, LDpred2-Inf and LDpred2 models are, in principle, special cases of the mixture model used in the SBayesR (although method implementations are different). In addition to traits with a highly polygenic genetic architecture, we have recently shown that SBayesR outperforms other methods for two less polygenic diseases, Alzheimer’s disease (49) (which includes the *APOE* locus which has a very large effect size) and amyotrophic lateral sclerosis (50) (for which there is evidence of greater importance of low MAF variants compared to SCZ(51)). The original SBayesR publication showed that in both simulations and applications to real data, the method performed well across a range of traits with different underlying genetic architectures. MegaPRS uses four different priors for the distribution of SNP effect, i.e.

Lasso, Ridge, BOLT-LMM, and BayesR (**Table 1**). It rescales SNP effects based on each of those priors and for each method selects the combination of parameters that maximises prediction in the tuning sample and then selects the best method amongst these. Hence, MegaPRS is a collection of the other methods and the SNP distribution selected varies depending on both tuning and target (**Table S11**). It selects BayesR 87% of the time when the tuning samples were large (otherwise BOLT-LMM) and selects Lasso 78% of the time when tuning samples were small. We implemented MegaPRS using the BLD-LDAK model recommended by the authors which assumes that the distribution of SNP effects depend on its allele frequency and functional annotation. While adding functional annotation to up or down weight SNPs is appealing, in practice there seemed to be no advantage in MegaPRS compared to LDpred2 and SBayesR that did not use functional annotations. Surprisingly, LDpred-funct method performed consistently less well than LDpred2-Inf, but this should be revisited as currently LDpred-funct is only available as a preprint (16).

Another study has compared 8 PGS methods for 8 disease/disorder traits (including MDD) and 3 continuous phenotypes comparing methods in two large community samples, the UK Biobank and the Twins Early Development Study (52). Consistent with our results, SBayesR attained a high prediction accuracy for MDD although they reported performance of SBayesR varied across traits. Since SBayesR expects effect size estimates and their standard errors to have properties consistent with the sample size and with the LD patterns imposed from an external reference panel, if GWAS summary statistics have non-ideal properties (perhaps resulting from meta-analysis errors or approximations) then SBayesR may not achieve converged solutions. SBayesR, in general, is more sensitive to any inconsistent properties between GWAS and LD reference samples than those methods that select hyper- parameters based on cross-validation in a tuning sample, such as LDpred2 (15). We note that the LDpred-funct preprint reported SBayesR to perform well across a range of quantitative and binary traits. A key advantage of SBayesR is that there is no need for the user to tune or select model or software parameters. Moreover, it does not need a tuning cohort to derive SNP effect weights but learns the genetic architecture from the properties of the GWAS results. Computationally it is also very efficient, using one CPU, it takes approximately 2 hours to generate SNP weights based on each discovery sample and predict into the left-out-cohort using a MCMC chain of 10,000 iterations (the computing time can be reduced by running a shorter chain since a negligible change in prediction accuracy was found after 4,000 iterations), which compares to PRS-CS: 40 hours using 5 CPUs, LDpred2: 5 hours using 15 CPUs, MegaPRS: 1 hours using 5 CPUs. Last, given that SBayesR uses only HapMap3 SNPs that are mostly well-imputed it should be possible to provide these SBayesR SNP weights as part of a GWAS pipeline to apply in external target samples.

All methods are compared using their default parameters settings. An optimum setting of each method could potentially increase the prediction accuracy. Most likely the optimum parameter settings are trait (genetic architecture) dependent(13). Here, we find that all methods that more formally model the genetic architecture than PC+T perform better than the PC+T, but there is little to choose between those methods. For application in psychiatric disorders, which are all highly polygenic traits, we particularly recommend LDpred2, MegaPRS and SBayesR which consistently rank high in all comparisons.

## Data Availability

The datasets stored in the Psychiatric Genomics Consortium central server follow strict guidelines with local ethics committee approval.

## Acknowledgements and Disclosures

We acknowledge funding from the National health and Medical Research Council (1173790,1078901,108788 (NRW),1113400 (NRW, PMV)) and the Australian Research Council (FL180100072 (PMV)).

This work would not have been possible without the contributions of the investigators who comprise the PGC-SCZ and PGC-MDD working group. For a full list of acknowledgments of all individual cohorts included in PGC-SCZ and PGC-MDD, please see the original publications. The PGC has received major funding from the US National Institute of Mental Health and the US National Institute of Drug Abuse (U01 MH109528 and U01 MH1095320). We thank the customers, research participants and employees of 23andMe for making this work possible. The study protocol used by 23andMe was approved by an external AAHRPP-accredited institutional review board.

The Münster cohort was funded by the German Research Foundation (DFG, grant FOR2107 DA1151/5-1 and DA1151/5-2 to U.D.; SFB-TRR58, Projects C09 and Z02 to U.D.) and the Interdisciplinary Center for Clinical Research (IZKF) of the medical faculty of Münster (grant Dan3/012/17 to U.D.).

Some data used in this study were obtained from dbGaP. dbGaP accession phs000021: funding support for the Genome-Wide Association of Schizophrenia Study was provided by the National Institute of Mental Health (R01 MH67257, R01 MH59588, R01 MH59571, R01 MH59565, R01 MH59587, R01 MH60870, R01 MH59566, R01 MH59586, R01 MH61675, R01 MH60879, R01 MH81800, U01 MH46276, U01 MH46289, U01 MH46318, U01 MH79469, and U01 MH79470), and the genotyping of samples was provided through the Genetic Association Information Network (GAIN). Samples and associated phenotype data for the Genome-Wide Association of Schizophrenia Study were provided by the Molecular Genetics of Schizophrenia Collaboration (principal investigator P. V. Gejman, Evanston Northwestern Healthcare (ENH) and Northwestern University, Evanston, IL, USA). dbGaP accession phs000196: this work used in part data from the NINDS dbGaP database from the CIDR: NGRC PARKINSON’S DISEASE STUDY. dbGaP accession phs000187: High-Density SNP Association Analysis of Melanoma: Case–Control and Outcomes Investigation. Research support to collect data and develop an application to support this project was provided by P50 CA093459, P50 CA097007, R01 ES011740, and R01 CA133996 from the NIH.

Statistical analyses were carried out on the Genetic Cluster Computer (http://www.geneticcluster.org) hosted by SURFsara and financially supported by the Netherlands Scientific Organization (NWO 480-05-003) along with a supplement from the Dutch Brain Foundation and the VU University Amsterdam.

The authors declare no competing interests.

The Schizophrenia Working Group of the Psychiatric Genomics Consortium is a collaborative co-author for this article. The individual authors are (affiliations are listed in the Supplement file) Stephan Ripke, Benjamin M. Neale, Aiden Corvin, James T. R. Walters, Kai-How Farh, Peter A. Holmans, Phil Lee, Brendan Bulik-Sullivan, David A. Collier, Hailiang Huang, Tune H. Pers, Ingrid Agartz, Esben Agerbo, Margot Albus, Madeline Alexander, Farooq Amin, Silviu A. Bacanu, Martin Begemann, Richard A Belliveau Jr, Judit Bene, Sarah E. Bergen, Elizabeth Bevilacqua, Tim B Bigdeli, Donald W. Black, Richard Bruggeman, Nancy G. Buccola, Randy L. Buckner, William Byerley, Wiepke Cahn, Guiqing Cai, Dominique Campion, Rita M. Cantor, Vaughan J. Carr, Noa Carrera, Stanley V. Catts, Kimberley D. Chambert, Raymond C. K. Chan, Ronald Y. L. Chen, Eric Y. H. Chen, Wei Cheng, Eric F. C. Cheung, Siow Ann Chong, C. Robert Cloninger, David Cohen, Nadine Cohen, Paul Cormican, Nick Craddock, James J. Crowley, Michael Davidson, Kenneth L. Davis, Franziska Degenhardt, Jurgen Del Favero, Ditte Demontis, Dimitris Dikeos, Timothy Dinan, Srdjan Djurovic, Gary Donohoe, Elodie Drapeau, Jubao Duan, Frank Dudbridge, Naser Durmishi, Peter Eichhammer, Johan Eriksson, Valentina Escott-Price, Laurent Essioux, Ayman H. Fanous, Martilias S. Farrell, Josef Frank, Lude Franke, Robert Freedman, Nelson B. Freimer, Marion Friedl, Joseph I. Friedman, Menachem Fromer, Giulio Genovese, Lyudmila Georgieva, Ina Giegling, Paola Giusti-Rodríguez, Stephanie Godard, Jacqueline I. Goldstein, Vera Golimbet, Srihari Gopal, Jacob Gratten, Lieuwe de Haan, Christian Hammer, Marian L. Hamshere, Mark Hansen, Thomas Hansen, Vahram Haroutunian, Annette M. Hartmann, Frans A. Henskens, Stefan Herms, Joel N. Hirschhorn, Per Hoffmann, Andrea Hofman, Mads V. Hollegaard, David M. Hougaard, Masashi Ikeda, Inge Joa, Antonio Julià, René S. Kahn, Luba Kalaydjieva, Sena Karachanak-Yankova, Juha Karjalainen, David Kavanagh, Matthew C. Keller, James L. Kennedy, Andrey Khrunin, Yunjung Kim, Janis Klovins, James A. Knowles, Bettina Konte, Vaidutis Kucinskas, Zita Ausrele Kucinskiene, Hana Kuzelova-Ptackova, Anna K. Kähler, Claudine Laurent, Jimmy Lee, S. Hong Lee, Sophie E. Legge, Bernard Lerer, Miaoxin Li, Tao Li, Kung-Yee Liang, Jeffrey Lieberman, Svetlana Limborska, Carmel M. Loughland, Jan Lubinski, Jouko Lönnqvist, Milan Macek, Patrik K. E. Magnusson, Brion S. Maher, Wolfgang Maier, Jacques Mallet, Sara Marsal, Manuel Mattheisen, Morten Mattingsdal, Robert W. McCarley, Colm McDonald, Andrew M. McIntosh, Sandra Meier, Carin J. Meijer, Bela Melegh, Ingrid Melle, Raquelle I. Mesholam- Gately, Andres Metspalu, Patricia T. Michie, Lili Milani, Vihra Milanova, Younes Mokrab, Derek W. Morris, Ole Mors, Kieran C. Murphy, Robin M. Murray, Inez Myin-Germeys, Bertram Müller-Myhsok, Mari Nelis, Igor Nenadic, Deborah A. Nertney, Gerald Nestadt, Kristin K. Nicodemus, Liene Nikitina-Zake, Laura Nisenbaum, Annelie Nordin, Eadbhard O’Callaghan, Colm O’Dushlaine, F. Anthony O’Neill, Sang-Yun Oh, Ann Olincy, Line Olsen, Jim Van Os, Psychosis Endophenotypes International Consortium, Christos Pantelis, George N. Papadimitriou, Sergi Papiol, Elena Parkhomenko, Michele T. Pato, Tiina Paunio, Milica Pejovic-Milovancevic, Diana O. Perkins, Olli Pietiläinen, Jonathan Pimm, Andrew J. Pocklington, John Powell, Alkes Price, Ann E. Pulver, Shaun M. Purcell, Digby Quested, Henrik B. Rasmussen, Abraham Reichenberg, Mark A. Reimers, Alexander L. Richards, Joshua L. Roffman, Panos Roussos, Douglas M. Ruderfer, Veikko Salomaa, Alan R. Sanders, Ulrich Schall, Christian R. Schubert, Thomas G. Schulze, Sibylle G. Schwab, Edward M. Scolnick, Rodney J. Scott, Larry J. Seidman, Jianxin Shi, Engilbert Sigurdsson, Teimuraz Silagadze, Jeremy M. Silverman, Kang Sim, Petr Slominsky, Jordan W. Smoller, Hon-Cheong So, Chris C. A. Spencer, Eli A. Stahl, Hreinn Stefansson, Stacy Steinberg, Elisabeth Stogmann, Richard E. Straub, Eric Strengman, Jana Strohmaier, T. Scott Stroup, Mythily Subramaniam, Jaana Suvisaari, Dragan M. Svrakic, Jin P. Szatkiewicz, Erik Söderman, Srinivas Thirumalai, Draga Toncheva, Sarah Tosato, Juha Veijola, John Waddington, Dermot Walsh, Dai Wang, Qiang Wang, Bradley T. Webb, Mark Weiser, Dieter B. Wildenauer, Nigel M. Williams, Stephanie Williams, Stephanie H. Witt, Aaron R. Wolen, Emily H. M. Wong, Brandon K. Wormley, Hualin Simon Xi, Clement C. Zai, Xuebin Zheng, Fritz Zimprich, Naomi R. Wray, Kari Stefansson, Peter M. Visscher, Wellcome Trust Case-Control Consortium, Rolf Adolfsson, Ole A. Andreassen, Douglas H. R. Blackwood, Elvira Bramon, Joseph D. Buxbaum, Anders D. Børglum, Sven Cichon, Ariel Darvasi, Enrico Domenici, Hannelore Ehrenreich, Tõnu Esko, Pablo V. Gejman, Michael Gill, Hugh Gurling, Christina M. Hultman, Nakao Iwata, Assen V. Jablensky, Erik G. Jönsson, Kenneth S. Kendler, George Kirov, Jo Knight, Todd Lencz, Douglas F. Levinson, Qingqin S. Li, Jianjun Liu, Anil K. Malhotra, Steven A. McCarroll, Andrew McQuillin, Jennifer L. Moran, Preben B. Mortensen, Bryan J. Mowry, Markus M. Nöthen, Roel A. Ophoff, Michael J. Owen, Aarno Palotie, Carlos N. Pato, Tracey L. Petryshen, Danielle Posthuma, Marcella Rietschel, Brien P. Riley, Dan Rujescu, Pak C. Sham, Pamela Sklar, David St Clair, Daniel R. Weinberger, Jens R. Wendland, Thomas Werge, Mark J. Daly, Patrick F. Sullivan & Michael B. O’Donovan

The Major Depressive Disorder Working Group of the Psychiatric Genomics Consortium is a collaborative co-author for this article. The individual authors are (affiliations are listed in the Supplement file) Naomi R Wray, Stephan Ripke, Manuel Mattheisen, Maciej Trzaskowski, Enda M Byrne, Abdel Abdellaoui, Mark J Adams, Esben Agerbo, Tracy M Air, Till F M Andlauer, Silviu-Alin Bacanu, Marie Bækvad-Hansen, Aartjan T F Beekman, Tim B Bigdeli, Elisabeth B Binder, Julien Bryois, Henriette N Buttenschøn, Jonas Bybjerg-Grauholm, Na Cai, Enrique Castelao, Jane Hvarregaard Christensen, Toni-Kim Clarke, Jonathan R I Coleman, Lucía Colodro-Conde, Baptiste Couvy-Duchesne, Nick Craddock, Gregory E Crawford, Gail Davies, Ian J Deary, Franziska Degenhardt, Eske M Derks, Nese Direk, Conor V Dolan, Erin C Dunn, Thalia C Eley, Valentina Escott-Price, Farnush Farhadi Hassan Kiadeh, Hilary K Finucane, Jerome C Foo, Andreas J Forstner, Josef Frank, Héléna A Gaspar, Michael Gill, Fernando S Goes, Scott D Gordon, Jakob Grove, Lynsey S Hall, Christine Søholm Hansen, Thomas F Hansen, Stefan Herms, Ian B Hickie, Per Hoffmann, Georg Homuth, Carsten Horn, Jouke-Jan Hottenga, David M Hougaard, David M Howard, Marcus Ising, Rick Jansen, Ian Jones, Lisa A Jones, Eric Jorgenson, James A Knowles, Isaac S Kohane, Julia Kraft, Warren W. Kretzschmar, Zoltán Kutalik, Yihan Li, Penelope A Lind, Donald J MacIntyre, Dean F MacKinnon, Robert M Maier, Wolfgang Maier, Jonathan Marchini, Hamdi Mbarek, Patrick McGrath, Peter McGuffin, Sarah E Medland, Divya Mehta, Christel M Middeldorp, Evelin Mihailov, Yuri Milaneschi, Lili Milani, Francis M Mondimore, Grant W Montgomery, Sara Mostafavi, Niamh Mullins, Matthias Nauck, Bernard Ng, Michel G Nivard, Dale R Nyholt, Paul F O’Reilly, Hogni Oskarsson, Michael J Owen, Jodie N Painter, Carsten Bøcker Pedersen, Marianne Giørtz Pedersen, Roseann E Peterson, Wouter J Peyrot, Giorgio Pistis, Danielle Posthuma, Jorge A Quiroz, Per Qvist, John P Rice, Brien P. Riley, Margarita Rivera, Saira Saeed Mirza, Robert Schoevers, Eva C Schulte, Ling Shen, Jianxin Shi, Stanley I Shyn, Engilbert Sigurdsson, Grant C B Sinnamon, Johannes H Smit, Daniel J Smith, Hreinn Stefansson, Stacy Steinberg, Fabian Streit, Jana Strohmaier, Katherine E Tansey, Henning Teismann, Alexander Teumer, Wesley Thompson, Pippa A Thomson, Thorgeir E Thorgeirsson, Matthew Traylor, Jens Treutlein, Vassily Trubetskoy, André G Uitterlinden, Daniel Umbricht, Sandra Van der Auwera, Albert M van Hemert, Alexander Viktorin, Peter M Visscher, Yunpeng Wang, Bradley T. Webb, Shantel Marie Weinsheimer, Jürgen Wellmann, Gonneke Willemsen, Stephanie H Witt, Yang Wu, Hualin S Xi, Jian Yang, Futao Zhang, Volker Arolt, Bernhard T Baune, Klaus Berger, Dorret I Boomsma, Sven Cichon, Udo Dannlowski, EJC de Geus, J Raymond DePaulo, Enrico Domenici, Katharina Domschke, Tõnu Esko, Hans J Grabe, Steven P Hamilton, Caroline Hayward, Andrew C Heath, Kenneth S Kendler, Stefan Kloiber, Glyn Lewis, Qingqin S Li, Susanne Lucae, Pamela AF Madden, Patrik K Magnusson, Nicholas G Martin, Andrew M McIntosh, Andres Metspalu, Ole Mors, Preben Bo Mortensen, Bertram Müller-Myhsok, Merete Nordentoft, Markus M Nöthen, Michael C O’Donovan, Sara A Paciga, Nancy L Pedersen

## Supplement 1

### Data

Schizophrenia GWAS summary statistics, were available from a total of 37 European ancestry cohorts reported in Pardiñas et al(1), comprising a total of 31K SCZ cases and 41K controls and 8M imputed SNPs. This included 34 cohorts from the PGC Schizophrenia (SCZ) Working group for which individual level genotype data were available. Detailed information about the cohorts is provided elsewhere(2) but is summarised in **Table S1**. PGS were calculated in each of the 30 PGC cohorts (target samples) using the GWAS discovery sample based on a meta-analysis of 37-2 = 35 cohorts i.e., the target sample was excluded from the discovery sample as well as a sample selected to be a tuning sample. Analyses were repeated using four different tuning samples, two of which were large (swe6:1094 cases and 1219 controls, gras: 1086 cases and 1232 controls) and two were small (lie2: 137 cases, 269 controls; msaf: 327 cases, 139 controls).

Major depression GWAS summary statistics from European ancestry studies comprised almost 13M imputed SNPs from 248K cases and 563K controls (3), which included data from the PGC Major Depressive Disorder (MDD) Working group (previously denoted as PGC29, but here MDD29). MDD29 includes data from 29 research study cohorts, described elsewhere (3–10) and summarised in **Table S2**. Individual level genotype data were available for 15K cases and 24K controls from 26 cohorts. We left one cohort out of those 26 cohorts in turn as the target sample, and then meta-analysed the remaining 28 samples with the other MDD GWAS summary statistics results to make the discovery samples. A cohort from Münster (3), not included in the discovery GWAS was used as the tuning sample (845 clinical defined MDD cases and 834 controls). Although the discovery sample meta-analyses include samples where the depression phenotype is self-reported rather than following a structured clinical interview, we refer to the prediction as MDD since the PGC target cohorts are of MDD cases and controls.

The datasets stored in the PGC central server follow strict guidelines with local ethics committee approval.

### Prediction methods

#### P-value based clumping and thresholding (PC+T)

In the PC+T method (also known as P+T or C+T)(11, 12) GWAS summary statistics are clumped to be approximately independent using a LD threshold, *r*^2^. From this quasi-independent genome-wide SNP list, SNPs are selected by thresholding on a pre-specified association p-value, Pt. We evaluated PC+T as implemented in Ricopili (13) as used in analyses of the (Psychiatric Genomics Consortia) which uses PLINK (14) to clump the SNP set using *r*^2^ = 0.1 within 500 kb windows, and Pt∈ (5e-08, 1e-06, 1e-04, 1e-03, 0.01, 0.05, 0.1, 0.2, 0.5, 1), where Pt =1 means that all SNPs from the LD-clumped list are included. In applications of PC+T it is common for results from the most associated Pt to be reported (including the application in the software PRSice (15) which uses a continuous Pt range), but this approach utilises information from the target cohort and hence introduces a form of winner’s curse. Here, the Pt threshold applied in target cohorts is the Pt threshold that maximised prediction in the tuning cohort.

#### SBLUP

SBLUP (16) is a method that re-scales the GWAS SNP effect estimates using an external LD reference panel to transform the ordinary least-squares estimates to approximate the best linear unbiased prediction (BLUP) solutions. This method assumes an infinitesimal model where SNP effects are drawn from a normal distribution. All genome-wide SNPs are used to build the PGS. Hence, for example, consider a genomic region with a single causal variant but with many SNPs in the region correlated with the causal variant and correlated with each other. In this case the SBLUP effect size estimate is “smeared” across the correlated SNPs, but with the total contribution to risk expected to represent the best estimate of the signal from the underlying causal variant. This method is implemented within the software package GCTA (17).

#### LDpred2 and LDpred2-Inf

LDpred2 (18) uses the GWAS summary statistics and LD information from the external LD reference sample to infer the posterior mean effect size of each SNP, conditioning on the SNP effect estimates of other correlated SNPs. This method assumes a point-normal prior on the distribution of SNP effects such that only a fraction of SNPs with non-zero estimated effects are selected for inclusion in the PGS. LDpred2 has three hyperparameters: the fractions of causal SNPs (*π*, but denoted *p* in the original paper), SNP-based heritability (*h*^2^_*g*_), and sparsity. We used the same parameter setting as in (18). The fractions of causal SNPs *π* values are equally spaced on log scale, i.e. π ∈ (0.00010, 0.00018, 0.00032, 0.00056, 0.00100, 0.00180, 0.00320, 0.00560, 0.01, 0.018, 0.032, 0.056, 0.1,0.18, 0.32, 0.56, 1). The values for *h*^2^_*g*_ are set at 0.70, 1 and 1.40 folds of the LDSC estimate. The sparsity choices are “true” or “false”. Normally, due to sampling variation, the SNPs in the subset with zero variance do not have exactly zero effect size; when sparsity is “true”, it forces those SNPs with exactly zero effects. The hyperparameters that maximise the prediction in the tuning sample are applied in the target sample; those values can differ between target cohorts even though the same tuning cohort is used, reflecting the properties of the discovery sample which may change with each left-out target sample. LDpred2-Inf is equivalent to SBLUP as the genetic architecture model assumes all SNPs have non-zero contribution of the phenotype variance. In software applications the results can differ because of the LD reference sample used and the assumptions for determining the LD window. The LD reference used in LDpred2 was the one provided on its website, which was calculated based on 362,320 UK Biobank individuals. Despite, the potential differences in the software applications, we observed a high concordance of results between SBLUP and LDPred-Inf (**Table S7**).

LDpred2 applied here used the grid-model. We did not include the auto-model (which does not need a tuning sample), because firstly, the LDPred2 paper (18) shows it has similar performance to the grid-model. Secondly, the LDPred2 software requires individual level genotype data of the LD reference to implement the auto-model which is not provided with the software whereas it does provide an LD matrix derived from individual level genotype data. The LDpred2 was run genome-wide, instead of per chromosome, since it attains higher prediction accuracy(18).

#### LDpred-funct

LDpred-funct (19) is an extension of the LDpred-Inf (SBLUP equivalent) model but leverages trait-specific functional enrichments relative to the baseline-LD model (20) to up/down-weight SNP effects. The functional annotations include coding, conserved, regulatory and LD-related annotation. In the baseline-LD model, the enrichment of each category is jointly calculated via stratified LD score regression (21). LDpred-funct has a non-infinitesimal model version, but besides the discovery and training samples, it needs the phenotype of the target samples to identify a parameter (the number of bins, *K*, in the original paper (22)). Given that this method is still under peer review and given that we wish to avoid parameter estimation in the target sample, we continued only with the infinitesimal model version.

#### MegaPRS

We applied the MegaPRS (23) software based on the BLD-LDAK model as recommended by the authors. The BLD-LDAK model assumes the expected per SNP heritability varies with its MAF, LD, and functional annotation, compared to other compared methods (e.g. SBLUP, LDpred-Inf) that assume the expected per SNP heritability is constant (24). Based on the estimated per SNP heritability, MegaPRS constructs PGS using four priors: Lasso, Ridge, BOLT-LMM, BayesR. Each of those priors has different hyperparameters. We used the same parameters as the original paper (23), which generates 100 Lasso models, 11 Ridge regression, 132 BOLT-LMM, and 84 BayesR models. For BayesR the genetic architecture parameters are the same as SBayesR, assuming 4 distributions of SNP effects, but determining the #_#_ proportions and their scaling factors through a grid search in the tuning cohort. See the MegaPRS paper for more details of these methods, Zhang *et al*.(23).

Following Zhang *et al*., we used 20K individuals with European ancestry from UK Biobank as the reference panel. The SNP annotation information used in the BLD-LDAK model were from ldak website (http://dougspeed.com/bldldak/).

#### Lassosum

Using GWAS summary statistics and a LD reference panel, Lassosum (25) constructs the PGS in a penalized regression framework. Lassosum is a deterministic method, and a convex optimization problem. It rescales the SNPs effect $ by minimizing 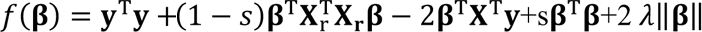, where **y** is the vector of phenotypes, -X_r_ is the genotype of LD reference; X is the genotype data of discovery sample, but this is not needed because X^T^y/N is the GWAS summary statistics that is known. Same as the original method paper, *s* set as 0.2, 0.5, 0.9, or 1. 0 are 20 values sequenced between 0.001 and 0.1 that equally spaced on the log-scale. The optimal hyperparameters for*s* and λ are identified in the tuning cohort. The current version of Lassosum cannot take a reference panel larger than 20K, and 5K is suggested (https://github.com/tshmak/lassosum). Hence, 5K unrelated UK Biobank individuals were randomly selected as the reference panel. We used only HapMap3 SNPs.

#### PRS-CS and PRS-CS-auto

PRS-CS (26) is also built under a Bayesian regression framework. Unlike LDpred2 which assumes a point-normal distribution as a prior, which is discrete, PRS-CS assumes a continuous shrinkage prior on the SNP effects. PRS-CS was implemented using the software default settings and with the LD reference panel provided with the PRS-CS software, which is computed using the 1000 Genomes samples and HapMap3 SNPs. In PRS-CS, for the global scaling parameter which is applied to all SNP effects *ϕ*, the search grid is *ϕ*^1/2^∈ (0.0001, 0.001, 0.01, 0.1, 1). The *ϕ* that produces the best predictive performance in a tuning data set is selected for use in the target sample. In PRS-CS-auto, *ϕ* is automatically learnt from GWAS summary statistics and no tunning sample is needed. *ψ* is a local marker-specific parameter which is drawn from the Gamma distribution, i.e. *ψ*_*j*_∼*Gamma*_*j*_) and δ_*j*_∼*Gamma*(b, 1). We used the default parameters proposed by the authors of *a* = 1 and *b* = 0.5.

#### SBayesR

SBayesR (27) is a method that re-scales the GWAS SNP effect estimates based on Bayesian multiple regression. SBayesR assumes that the standardised SNP effects are drawn from a mixture of C=4 zero-mean normal distributions with different variances (one of the variances is zero, with a probability of π1), indicating that only a fraction of SNPs (1-π1) have non-zero estimated effects which contribute to the phenotype. Moreover, the contributions of SNPs in different distributions differ because of different variances. Here, we evaluated SBayesR in the default setting. The scaling factor > for the variance of each mixture component are set as 0, 0.01, 0.1, and 1 in this order. The banded LD matrix was downloaded from GCTB website (https://cnsgenomics.com/software/gctb/~Download), which was built based on the HapMap3 SNPs of randomly selected and unrelated 10K UK Biobank individuals. The windows size used to estimate the LD is 3cM, which is the same as LDpred2. Whereas LDpred2 estimates π from a tuning sample, SBayesR estimates π from the GWAS discovery sample, so no tuning sample is needed. LDpred2 has an auto version which does need the tuning sample, but it requires individual level genotype data of the LD reference which is not provided with the software whereas it does provide an LD matrix derived from individual level genotype data.

### AUC vs variance explained on the liability scale

Although covariates were not included when calculating AUC the impact is small. For example, for SCZ the maximum median variance in liability was for MegaPRS at 9.2%. Assuming lifetime risk of SCZ of 0.01 the AUC expected from normal distribution theory(4) (see pseudo-code section) is 0.722, compared to the mean reported of 0.731. For MDD the maximum median variance in liability was for SBayesR at 3.5%. Assuming a lifetime risk of the expected AUC is 0.596 compared to the mean reported of 0.599. The AUC and variance in liability from the model including 6 principal components and PGS in the regression is in Table S3 and Table S5.

## Table titles and legends

**Figure S1.**
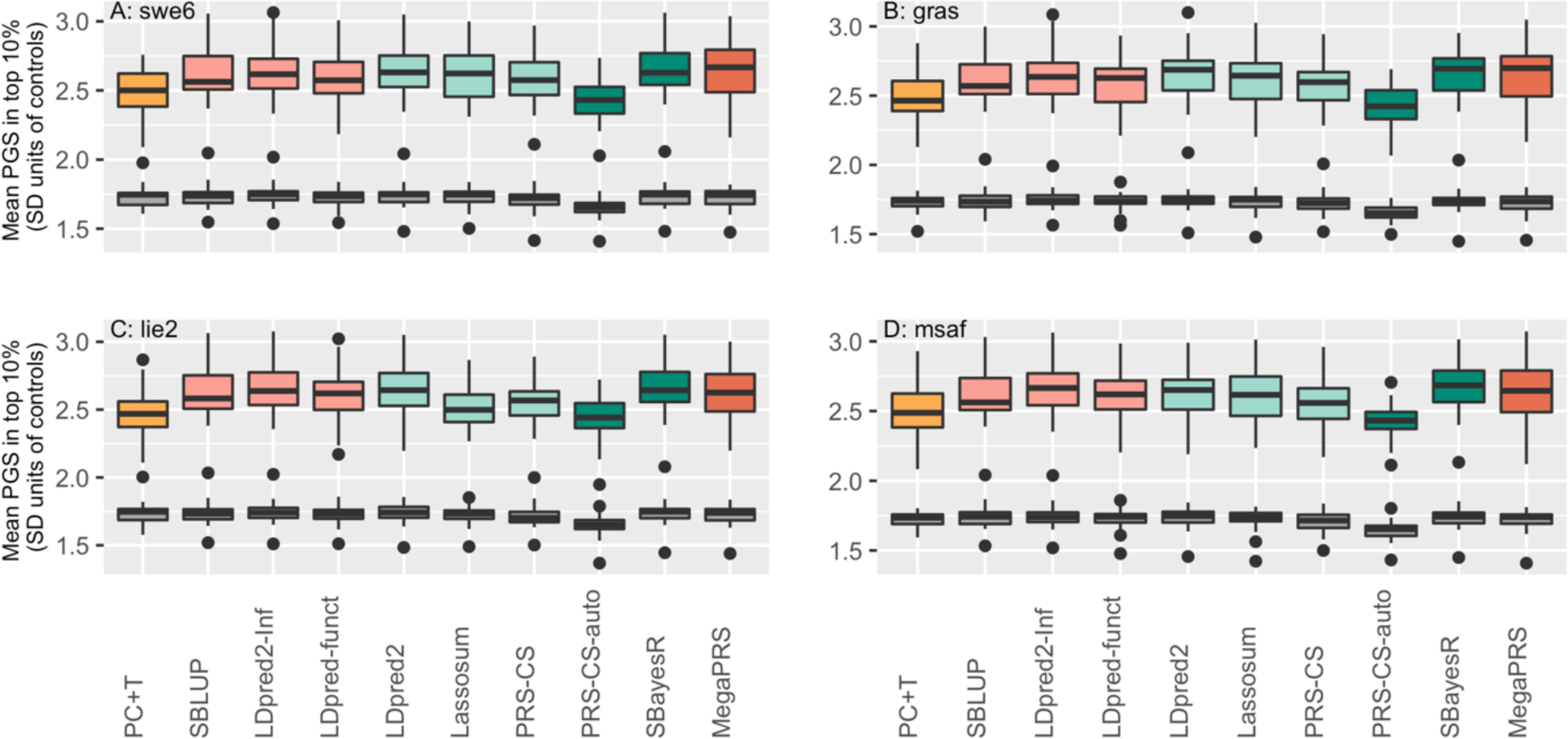
PGS in top 10% of SCZ cases and controls. The mean of the PGS for the top 10% cases (colored boxes) and for the top 10% of controls (grey boxes) in PGS standard deviation (SD) unit scale. The controls have mean PGS of zero and SD of 1. Subfigures are the results using different tuning cohorts. Since the PGS are normally distributed, as expected the mean PGS for controls in the top 10% PGS is ∼1.75 SD units, whereas the top 10% of cases have mean value of 2.65 control sample SD units using SBayesR. These mean values of the top 10% in cases equate to expectations from the population of the top 1.1% SCZ.

**Figure S2.**
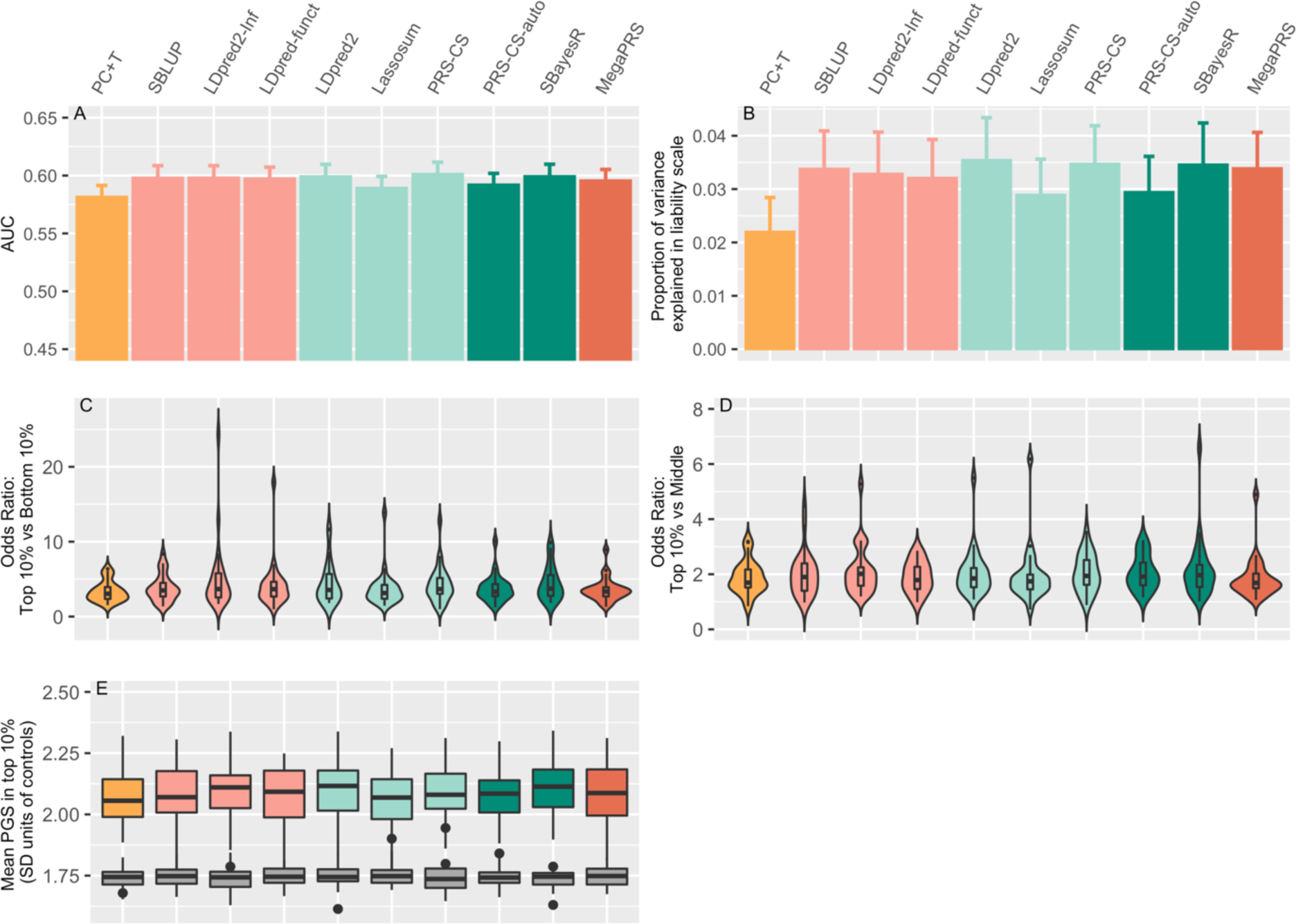
Prediction of MDD case/control status using different PGS methods. A) The area under curve (AUC) statistic. The AUC is a measure for the prediction accuracy, which indicates the probability that a case ranks higher than a control. The predictors were constructed from GWAS summary statistics of UK Biobank(4, 28), 23andMe(5), GERA(29), iPSYCH (7), deCODE (8), GenScotland (9, 10), PGC- MDD29 excluding the target cohort. The target cohorts comprised 26 of the 29 cohorts in MDD29. A cohort from Münster (845 clinical defined MDD cases and 834 controls), not included in the MDD29, was used as the tuning sample. Each bar reflects the median AUC across 26 target cohorts, the whiskers show the 95% confidence interval for comparing medians. B) The proportion of variance explained by PGS on the scale of liability, assuming a population lifetime risk of 15%. C) The odds ratio when considering the odds of being a case comparing the top 10% vs bottom 10% of PGS. D) The odds ratio when considering the odds of being a case comparing the top 10% vs those in the middle of the PGS distribution, calculated as the averaged odds ratio of the top 10% ranked on PGS relative to the 5^th^ decile and 6^th^ decile. E) The mean of the PGS for the top 10% cases (coloured boxes) and for the top 10% of controls (grey boxes) in PGS standard deviation (SD) unit scale so that controls have mean PGS of zero and SD of 1. Since the PGS are normally distributed, as expected the mean PGS for controls in the top 10% PGS is ∼1.75 SD units, whereas the top 10% of cases have mean value of 2.10 control sample SD units for MDD cases, using SBayesR. These mean values of the top 10% in cases equate to expectations from the population of the top 4.7% for MDD.

**Figure S3.**
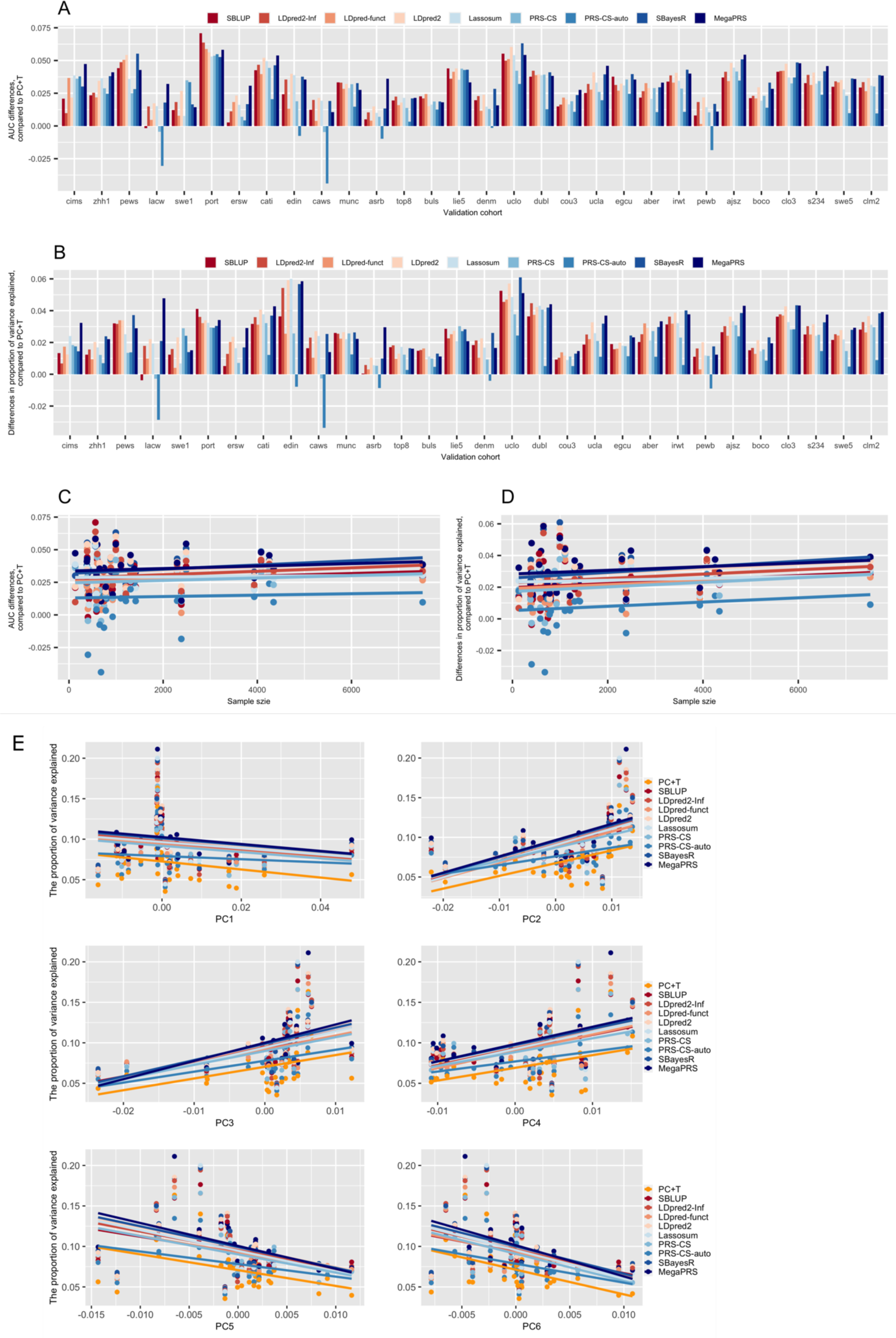
Individual SCZ cohort results and relationship with potential confounders. The area under the curve (A and C panels) and the proportion of variance explained by PGS on the liability scale (B and D panels) of schizophrenia predicted by different PGS methods in each of target cohorts, compared to PC+T method. x-axis of A and B are the target cohorts ordered by sample size, increasing from left (Ncases = 71, Ncontrols =69) to right (Ncases = 3466, Ncontrols =4297). x-axis of C and D are the sample sizes of each target cohorts. The lines in C and D are the regression lines of y and x by each method. For each method, when regressing AUC difference on the sample size of the target cohort, the p-values are all larger than 0.05. Similarly, the p-values of regressing the proportion of variance explained by PGS on the sample size are larger than 0.05. E) The proportion of variance explained on the liability scale against first 6 principal components (PCs), which were estimated from directly genotyped SNPs shared across cohorts. The x-axis is the mean value of the PC in the cohort. The regression p-values were: PC1: 0.25-0.56, PC2: 0.001-0.004, PC3: 0.014-0.052, PC4: 0.004-0.024, PC5: 0.016-0.049, PC6: 0.009-0.056, with the range reflecting different methods. Using the 23 European cohorts collected in a single country, we found in regression of each PC on latitude, longitude and SNP-array (Affymetrix, Illumina-nonOmni, Illumina Omi) the following significant associations (P<0.01): PC1: latitude & Array, PC2: longitude, PC3: latitude & longitude, PC4: Array, PC5: latitude & array, recognising that latitude and longitude could represent phenotype as well as genetic ancestry differences.

**Figure S4.**
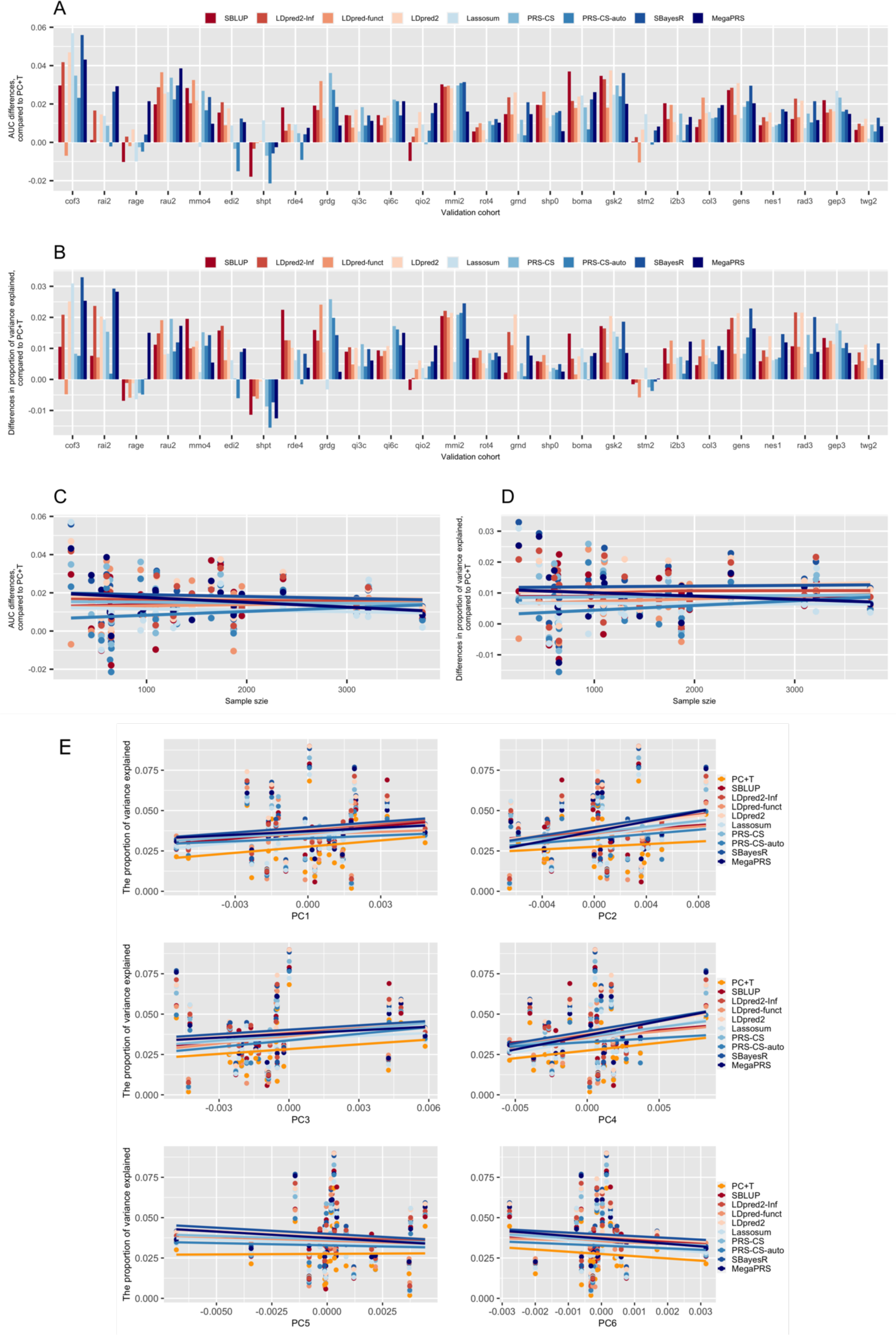
Individual MDD cohort results and relationship with potential confounders. The area under the curve (A and C panels) and the proportion of variance explained by PGS on the liability scale (B and D panels) of major depression predicted by different PGS methods in each of target cohorts, compared to PC+T method. x-axis of A and B are the target cohorts ordered by sample size, increasing from left (Ncases = 120, Ncontrols =126) to right (Ncases = 1,097, Ncontrols =2,663). x-axis of C and D are the sample sizes of each target cohorts. The lines in C and D are the regression lines of y and x for each method. For each method, when regressing AUC difference on the sample size of the target cohort, the p-values are all larger than 0.05. Similarly, the P-values of regressing the proportion of variance explained by PGS on the sample size are larger than 0.05. E) The proportion of variance explained on the liability scale against first 6 principal components (PCs), which were estimated from directly genotyped SNPs shared across cohorts. The x-axis is the mean value of the PC in the cohort. The regression p-values were: PC1: 0.39-0.76, PC2: 0.09-0.65, PC3: 0.28-0.64, PC4: 0.16-0.68, PC5: 0.62-0.96, PC6: 0.59-0.85, with the range reflecting different methods. Using the 15 European cohorts collected in a single country, we found in regression of each PC on latitude, longitude and SNP-array (Affymetrix, Illumina-nonOmni, Illumina Omi) the following significant associations (P<0.01): PC1: latitude, PC2: longitude, recognising that latitude and longitude could represent phenotype as well as genetic ancestry differences.

**Figure S5.**
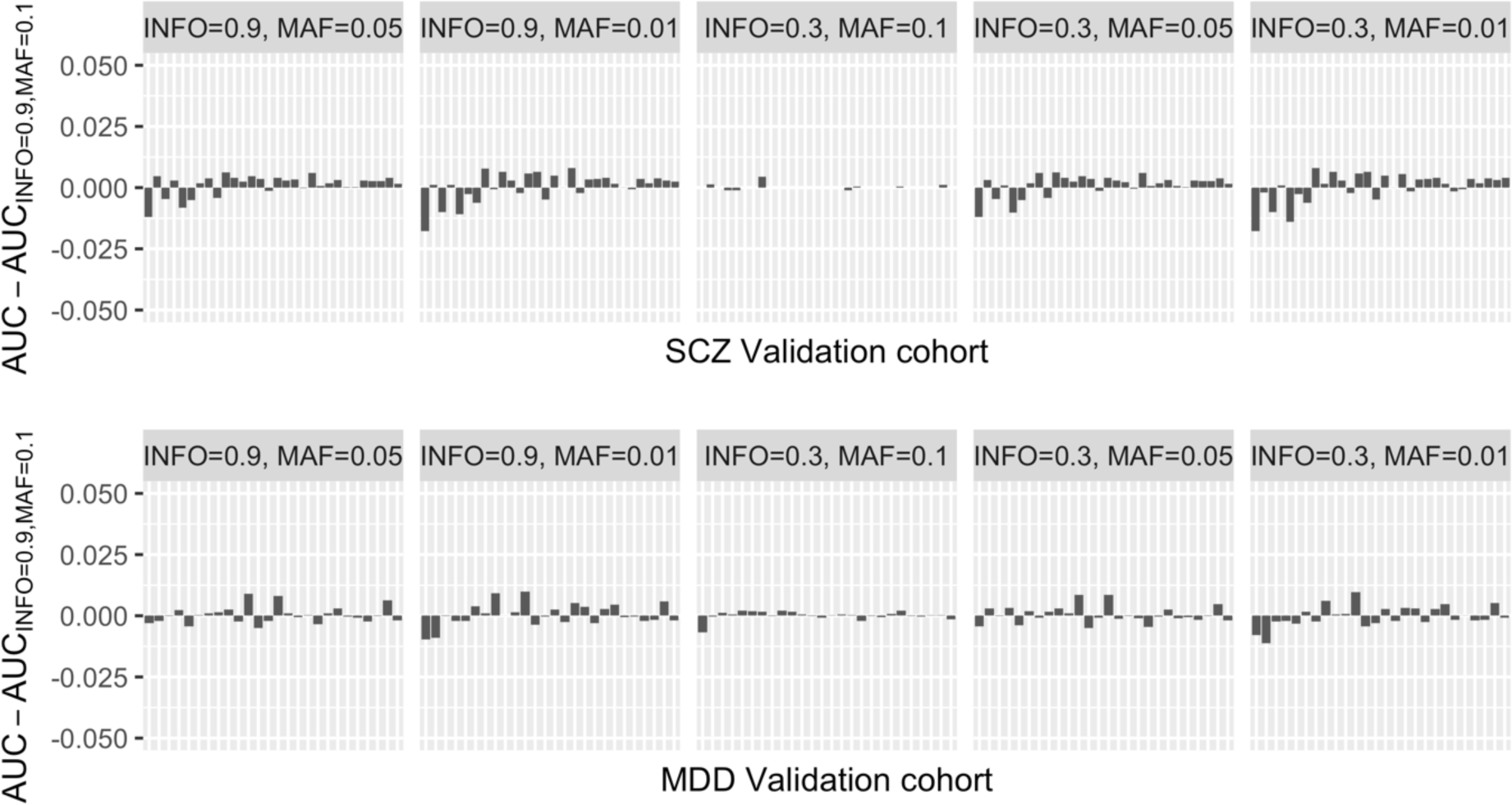
Sensitivity analysis: INFO score and MAF. Differences in AUC of SBayesR when using different quality control thresholds. The different bars refer to different target cohorts ordered by its sample size.

**Figure S6.**
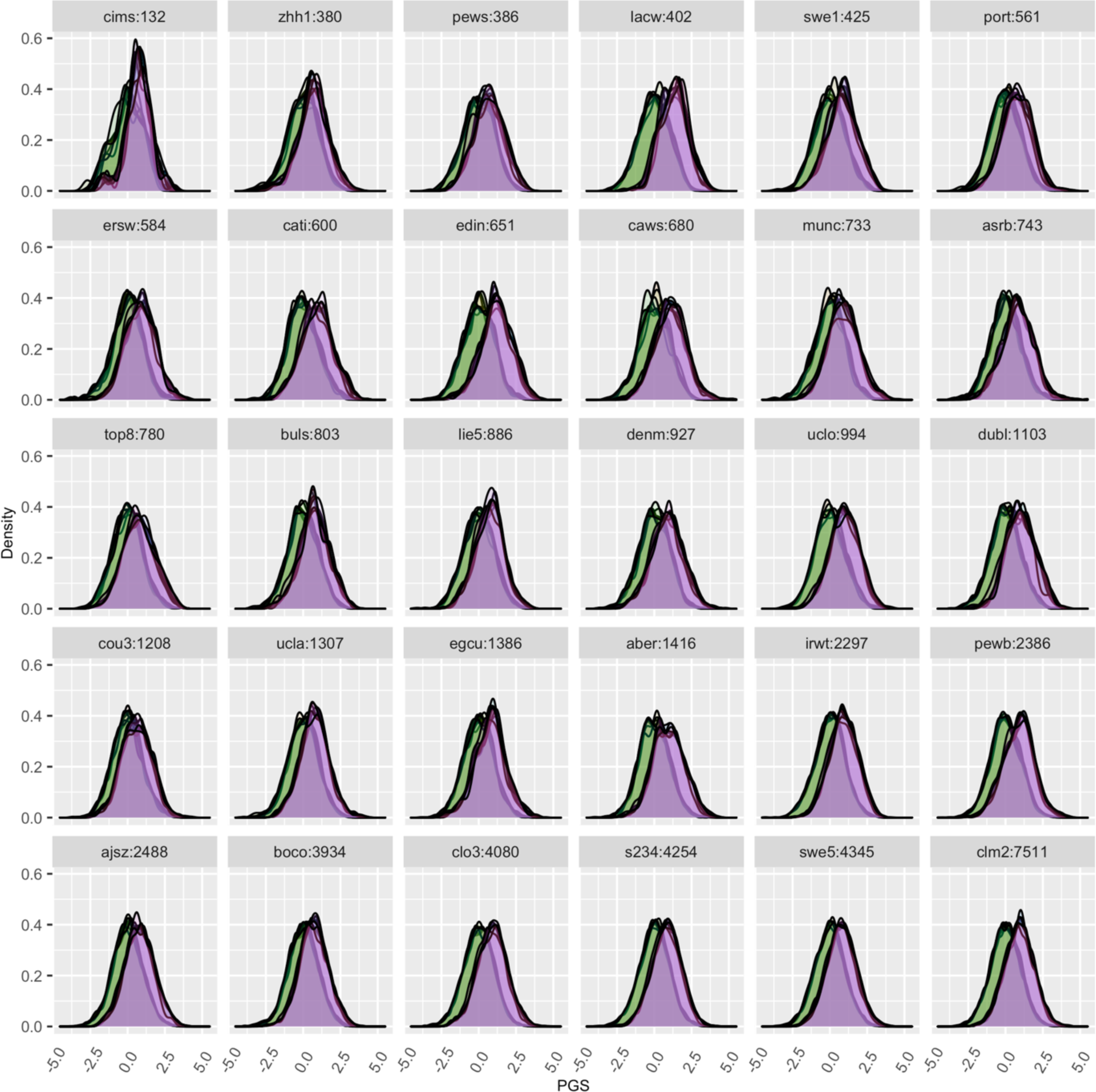
PGS densities of SCZ cases and controls in each target cohort ordered by sample size. Light green shows the PGS density of controls predicted by different methods. Light purple shows the PGS density of cases predicted by different methods. The PGS were scaled to SD units of controls. Thus, the mean and variance of PGS in controls are zero and one, respectively. The mean and variance of PGS in cases are in Table S3 (SD of PGS of cases (SD units of controls)).

**Figure S7.**
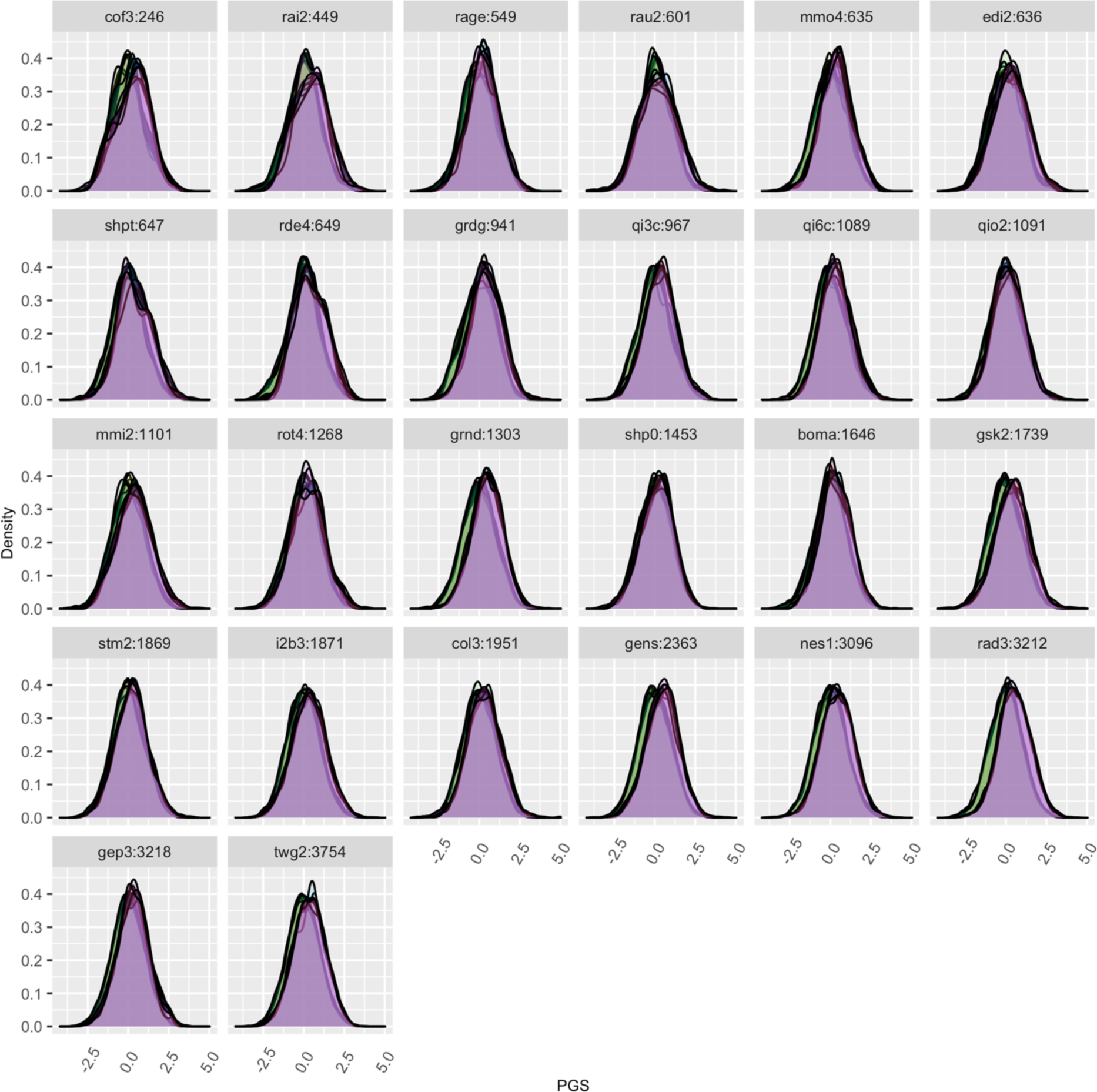
PGS densities of MDD cases and controls each target cohort ordered by sample size. Light green shows the PGS density of controls predicted by different methods. Light purple shows the PGS density of cases predicted by different methods. The PGS were scaled to SD units of controls. Thus, the mean and variance of PGS in controls are zero and one, respectively. The mean and variance of PGS in cases are in Table S5 (SD of PGS of cases (SD units of controls)).

**Figure S8.**
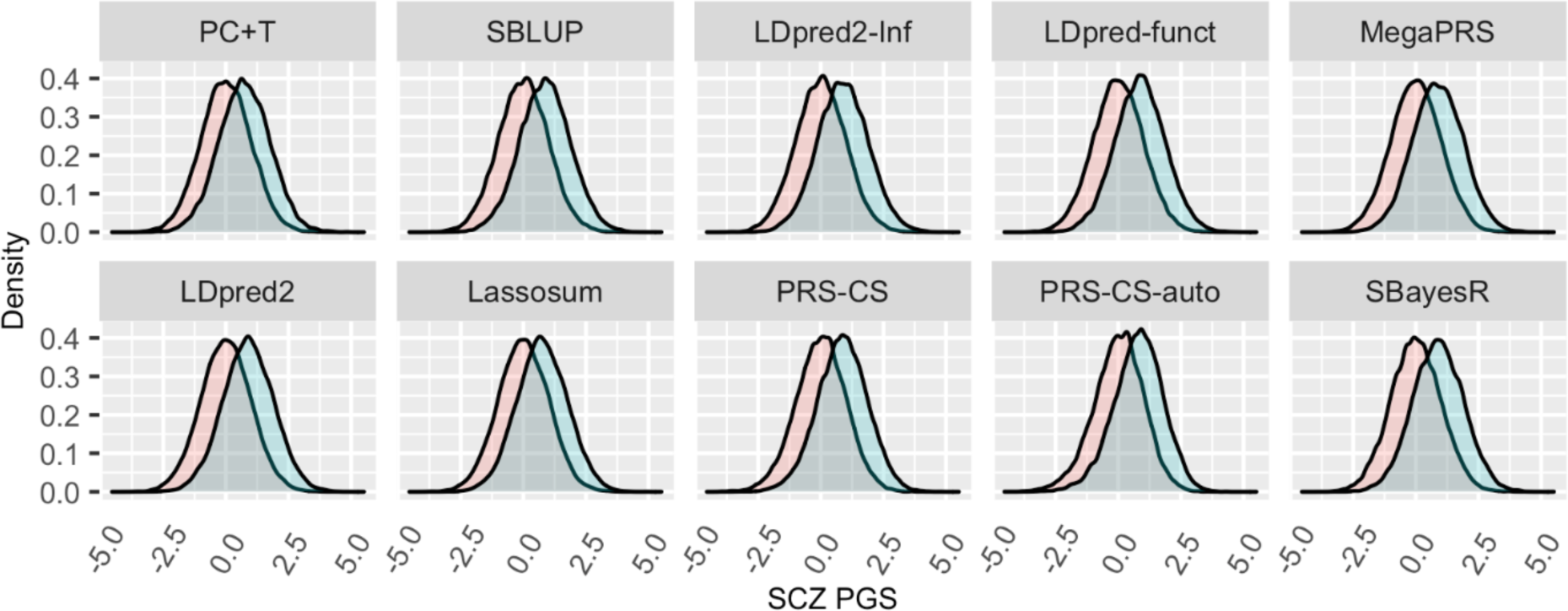
PGS densities of SCZ cases and controls estimated by different methods across the target cohorts. The mean PGS of cases is, on average, 0.85 standard deviation units (calculated in controls) and refer to Table S3 for estimate of each method.

**Figure S9.**
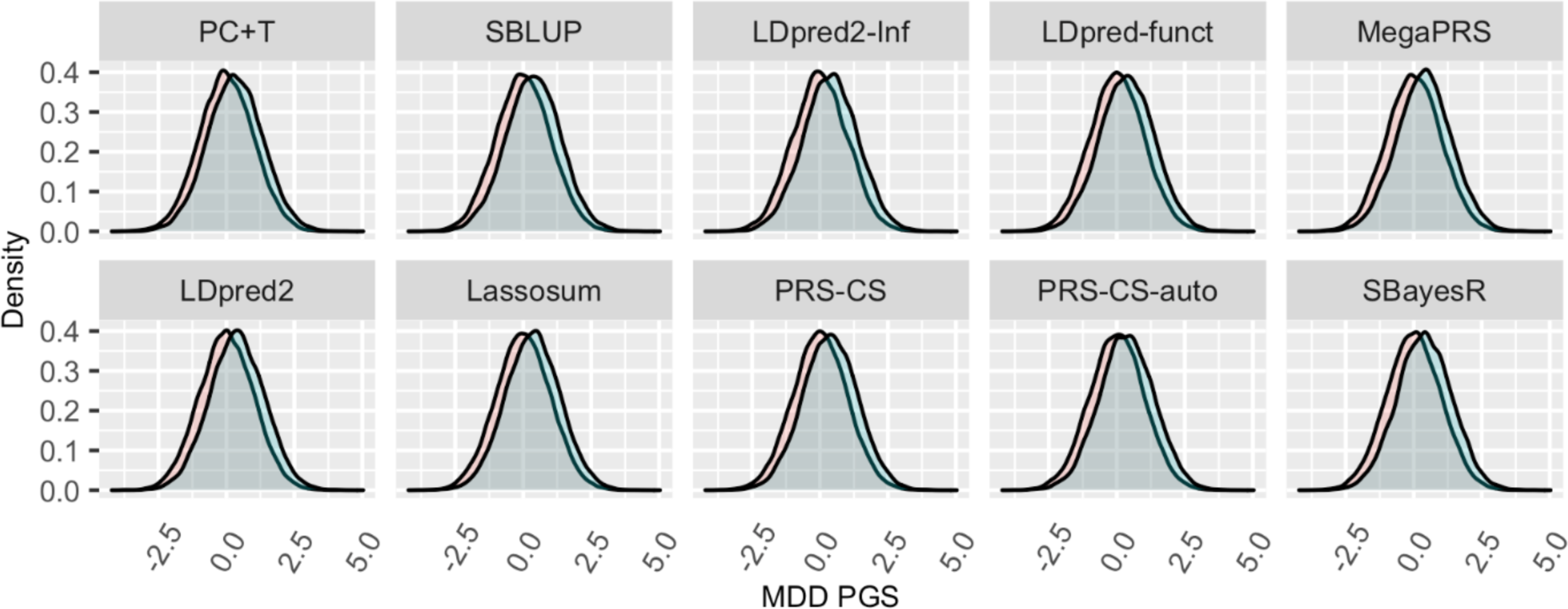
PGS densities of MDD cases and controls estimated by different methods across the target cohort. The mean PGS of cases is, on average, 0.34 standard deviation units (calculated in controls) and refer to Table S5 for estimate of each method.

### Pseudo code

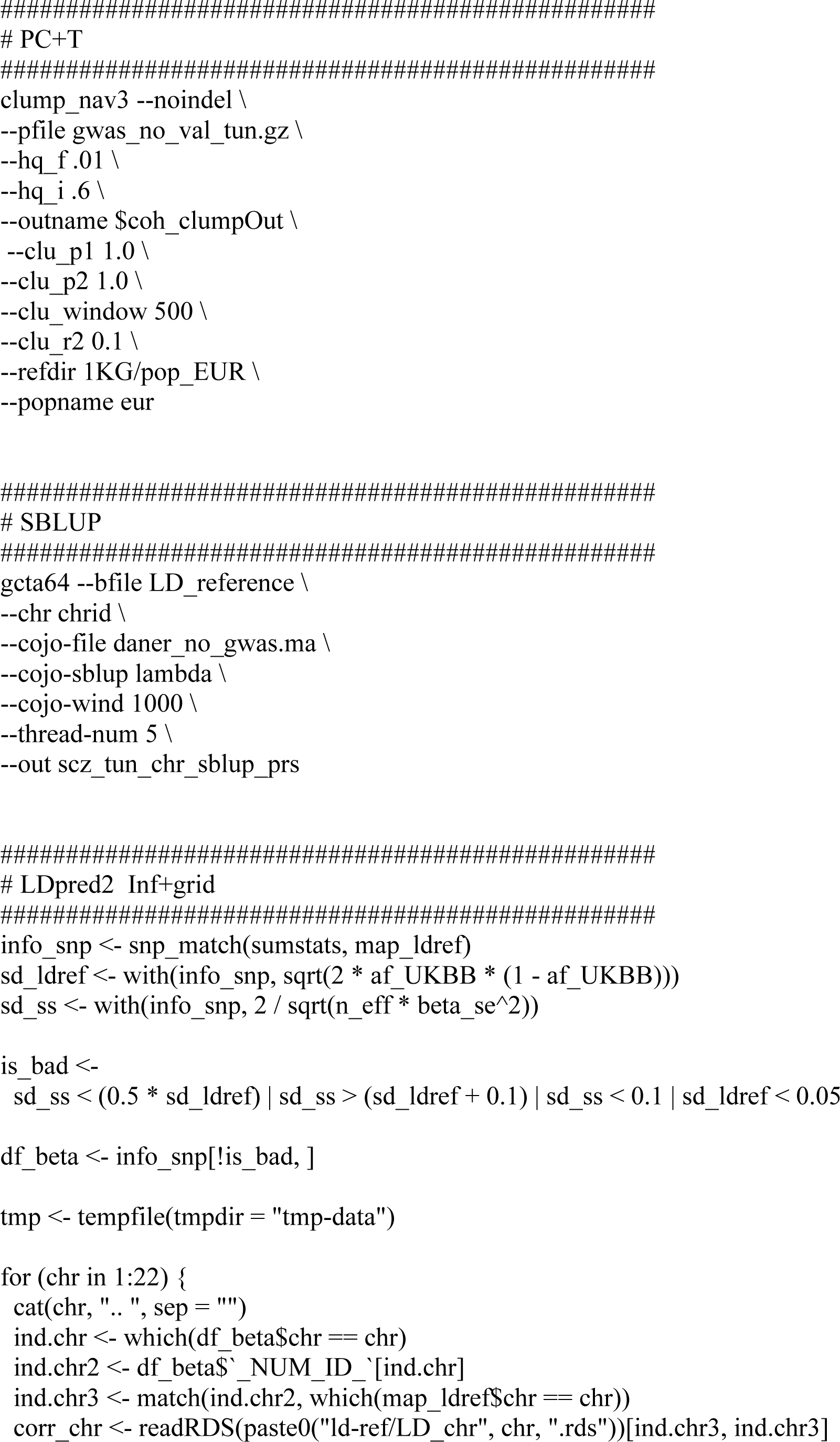
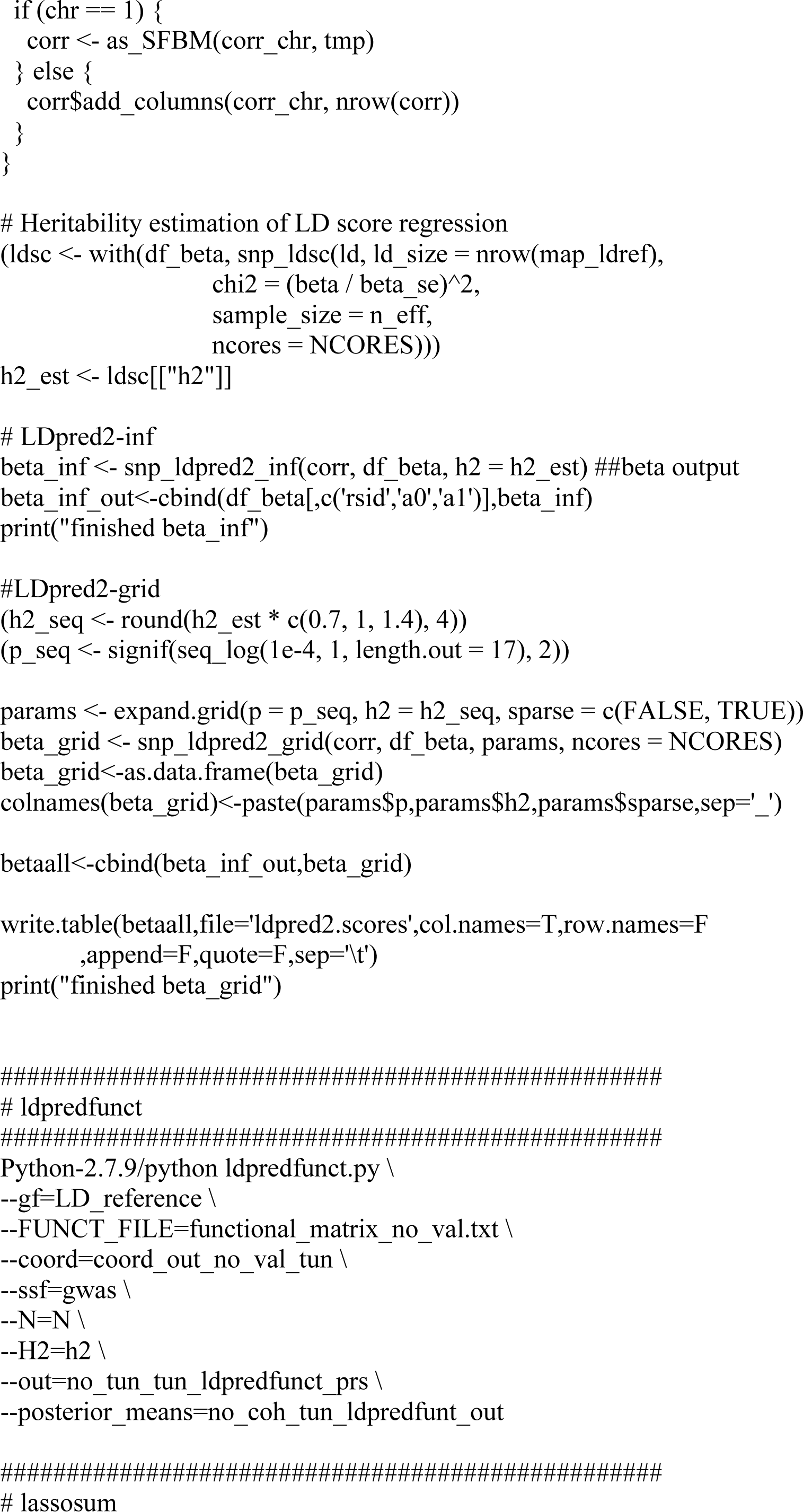
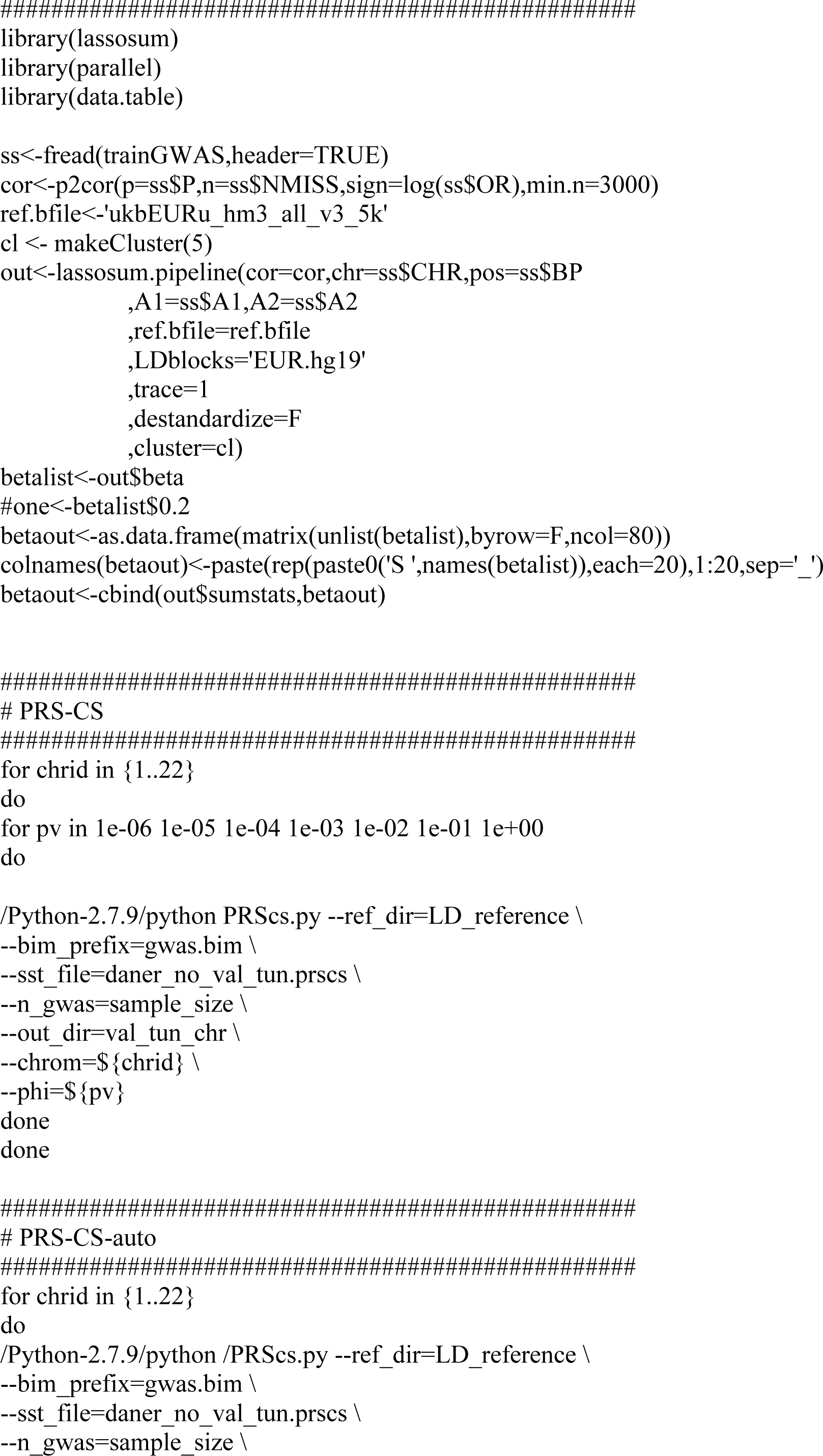
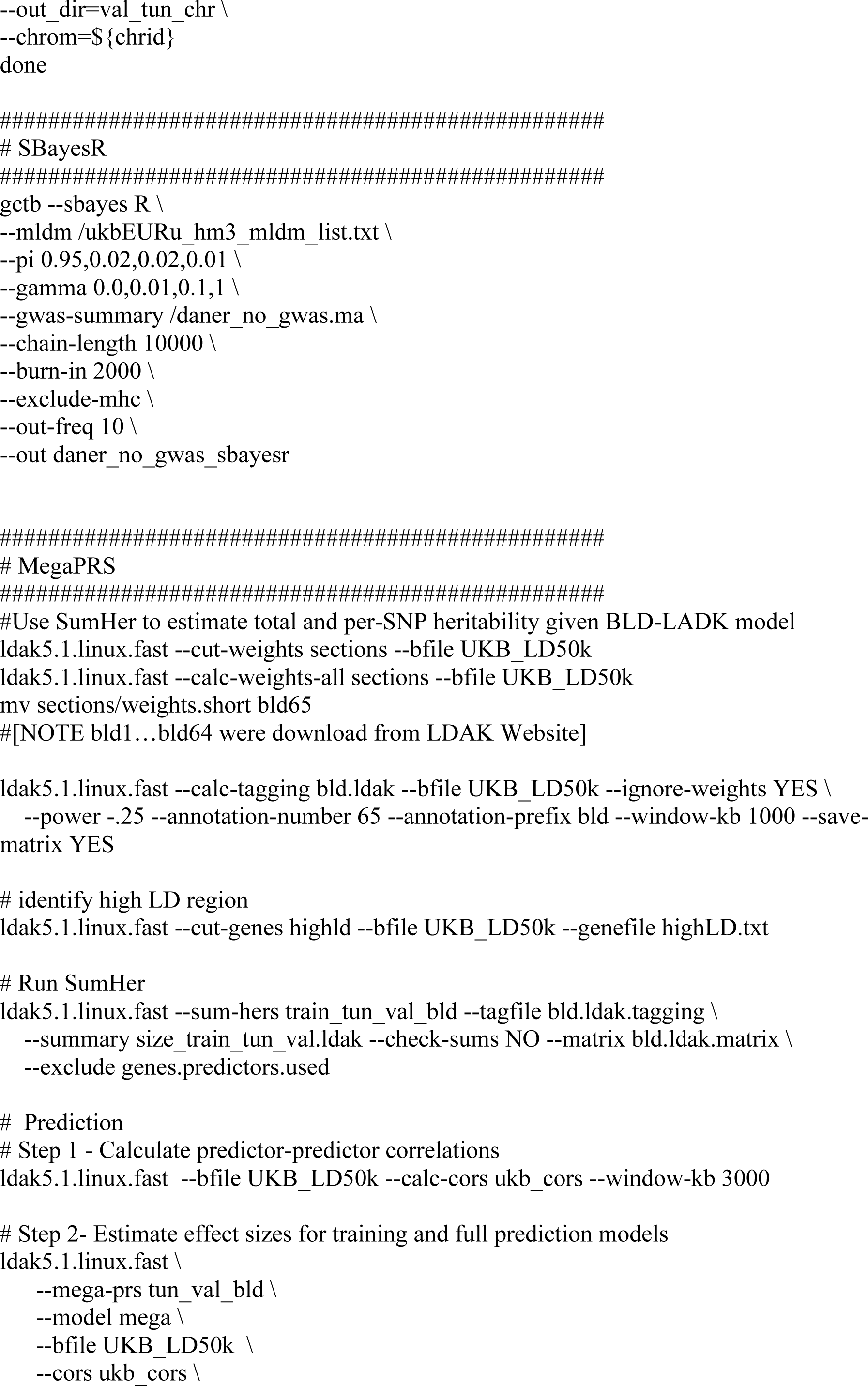
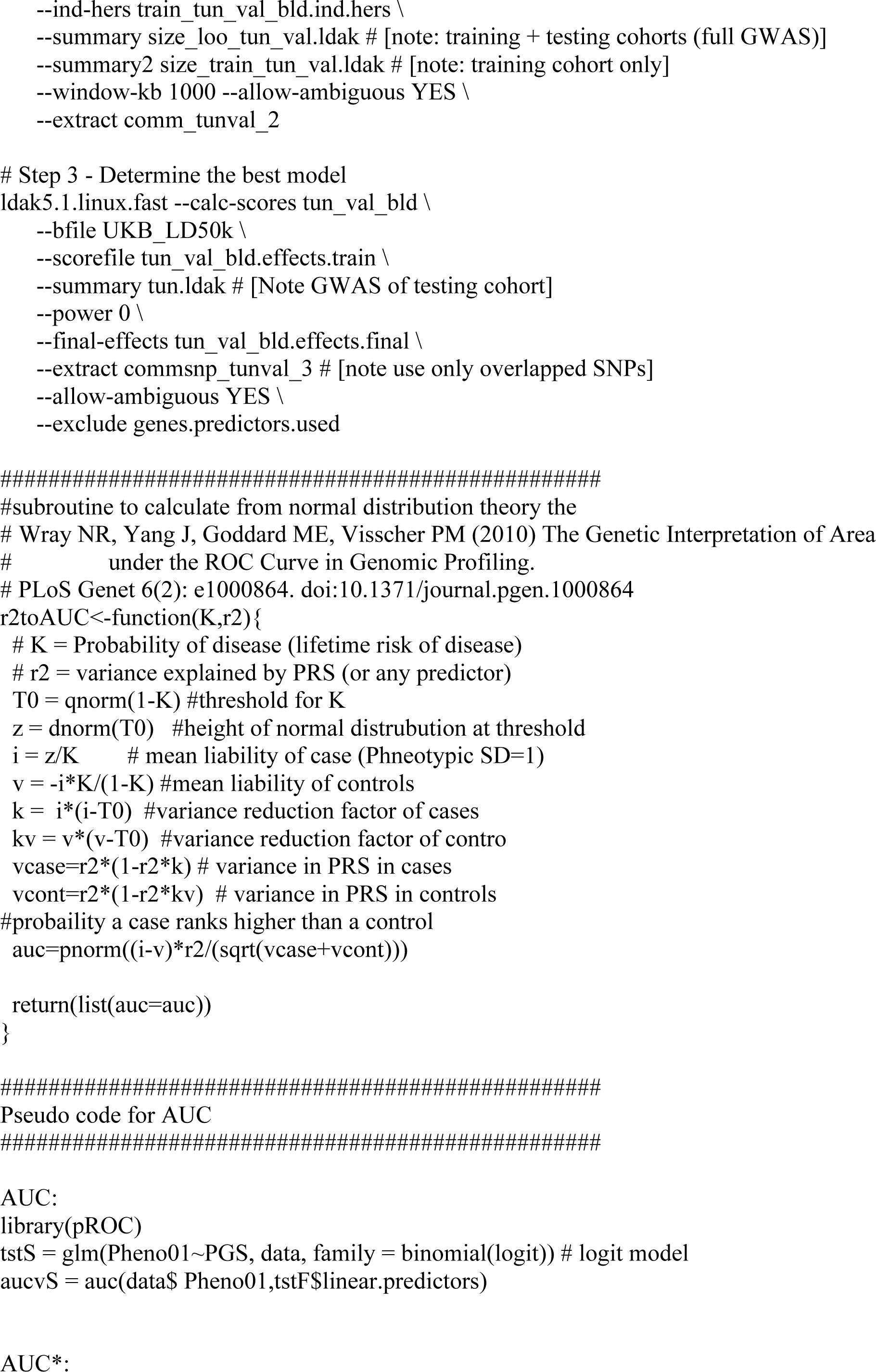
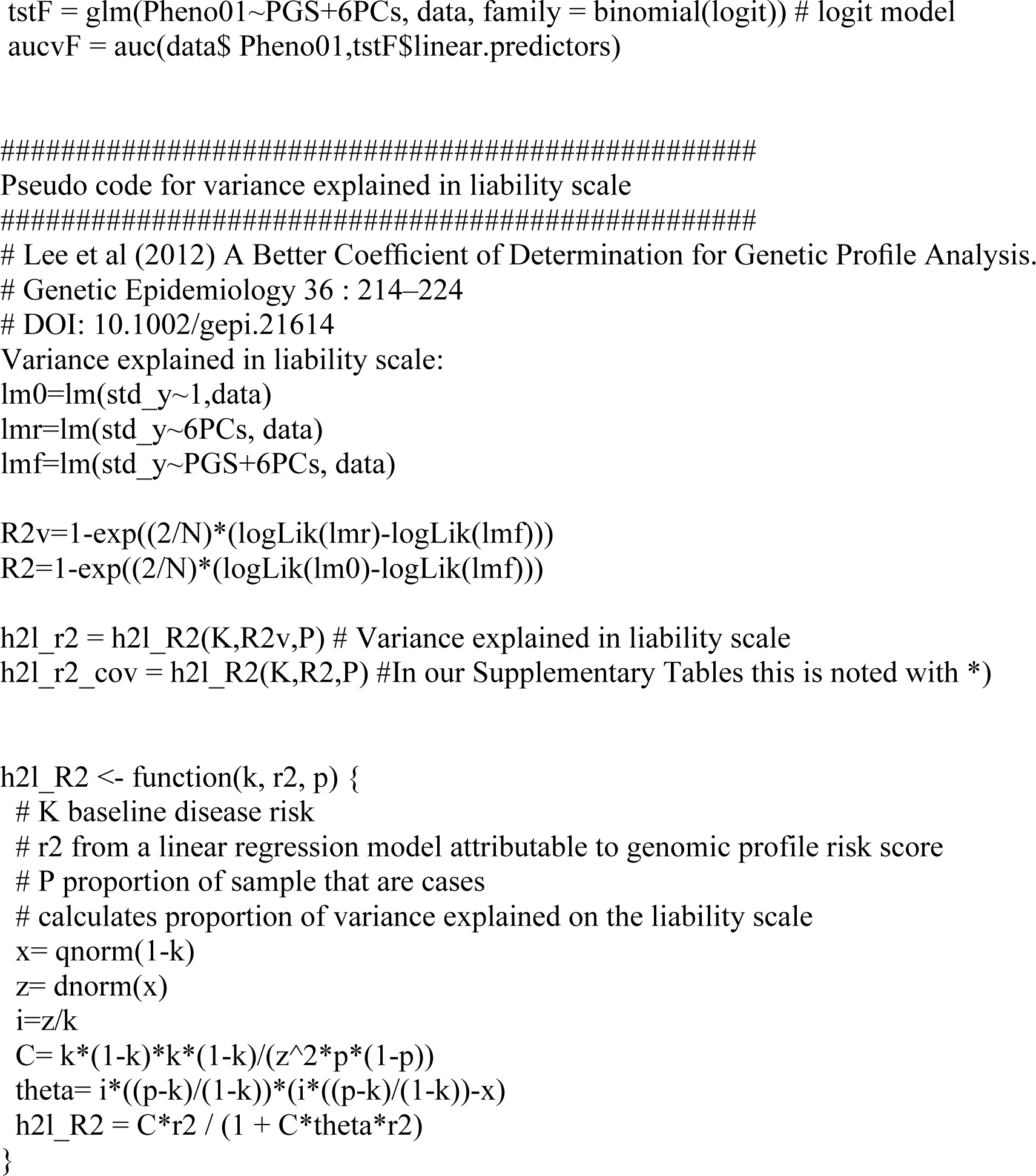

### Schizophrenia Working Group of the Psychiatric Genomics Consortium

Stephan Ripke^1,2^, Benjamin M. Neale^1,2,3,4^, Aiden Corvin^5^, James T. R. Walters^6^, Kai-How Farh^1^, Peter A. Holmans^6,7^, Phil Lee^1,2,4^, Brendan Bulik-Sullivan^1,2^, David A. Collier^8,9^, Hailiang Huang^1,3^, Tune H. Pers^3,10,11^, Ingrid Agartz^12,13,14^, Esben Agerbo^15,16,17^, Margot Albus^18^, Madeline Alexander^19^, Farooq Amin^20,21^, Silviu A. Bacanu^22^, Martin Begemann^23^, Richard A Belliveau Jr^2^, Judit Bene^24,25^, Sarah E. Bergen ^2,26^, Elizabeth Bevilacqua^2^, Tim B Bigdeli ^22^, Donald W. Black^27^, Richard Bruggeman^28^, Nancy G. Buccola^29^, Randy L. Buckner^30,31,32^, William Byerley^33^, Wiepke Cahn^34^, Guiqing Cai^35,36^, Dominique Campion^37^, Rita M. Cantor^38^, Vaughan J. Carr^39,40^, Noa Carrera^6^, Stanley V. Catts^39,41^, Kimberley D. Chambert^2^, Raymond C. K. Chan^42^, Ronald Y. L. Chen^43^, Eric Y. H. Chen^44^, Wei Cheng^45^, Eric F. C. Cheung^46^, Siow Ann Chong^47^, C. Robert Cloninger^48^, David Cohen^49^, Nadine Cohen^50^, Paul Cormican^5^, Nick Craddock^6,7^, James J. Crowley^51^, Michael Davidson^54^, Kenneth L. Davis^36^, Franziska Degenhardt^55,56^, Jurgen Del Favero^57^, Ditte Demontis^17,58,59^, Dimitris Dikeos^60^, Timothy Dinan^61^, Srdjan Djurovic^14,62^, Gary Donohoe^5,63^, Elodie Drapeau^36^, Jubao Duan^64,65^, Frank Dudbridge^66^, Naser Durmishi^67^, Peter Eichhammer^68^, Johan Eriksson^69,70,71^, Valentina Escott-Price^6^, Laurent Essioux^72^, Ayman H. Fanous^73,74,75,76^, Martilias S. Farrell^51^, Josef Frank^77^, Lude Franke^78^, Robert Freedman^79^, Nelson B. Freimer^80^, Marion Friedl^81^, Joseph I. Friedman^36^, Menachem Fromer^1,2,4,82^, Giulio Genovese^2^, Lyudmila Georgieva^6^, Ina Giegling^81,83^, Paola Giusti-Rodríguez^51^, Stephanie Godard^84^, Jacqueline I. Goldstein^1,3^, Vera Golimbet^85^, Srihari Gopal^86^, Jacob Gratten^87^, Lieuwe de Haan^88^, Christian Hammer^23^, Marian L. Hamshere^6^, Mark Hansen^89^, Thomas Hansen^17,90^, Vahram Haroutunian^36,91,92^, Annette M. Hartmann^81^, Frans A. Henskens^39,93,94^, Stefan Herms^55,56,95^, Joel N. Hirschhorn^3,11,96^, Per Hoffmann^55,56,95^, Andrea Hofman^55,56^, Mads V. Hollegaard^97^, David M. Hougaard^97^, Masashi Ikeda^98^, Inge Joa^99^, Antonio Julià^100^, René S. Kahn^101^, Luba Kalaydjieva^102,103^, Sena Karachanak-Yankova^104^, Juha Karjalainen^78^, David Kavanagh^6^, Matthew C. Keller^105^, James L. Kennedy^106,107,108^, Andrey Khrunin^109^, Yunjung Kim^51^, Janis Klovins^110^, James A. Knowles^111^, Bettina Konte^81^, Vaidutis Kucinskas^112^, Zita Ausrele Kucinskiene^112^, Hana Kuzelova-Ptackova^113,114^, Anna K. Kähler^26^, Claudine Laurent^19,115^, Jimmy Lee^47,116^, S. Hong Lee^87^, Sophie E. Legge^6^, Bernard Lerer^117^, Miaoxin Li^118^, Tao Li^119^, Kung-Yee Liang^120^, Jeffrey Lieberman^121^, Svetlana Limborska^109^, Carmel M. Loughland^39,122^, Jan Lubinski^123^, Jouko Lönnqvist^124^, Milan Macek^113,114^, Patrik K. E. Magnusson^26^, Brion S. Maher^125^, Wolfgang Maier^126^, Jacques Mallet^127^, Sara Marsal^100^, Manuel Mattheisen^17,58,59,128^, Morten Mattingsdal^14,129^, Robert W. McCarley^130,131^, Colm McDonald^132^, Andrew M. McIntosh^133,134^, Sandra Meier^77^, Carin J. Meijer^88^, Bela Melegh^24,25^, Ingrid Melle^14,135^, Raquelle I. Mesholam-Gately^130,136^, Andres Metspalu^137^, Patricia T. Michie^39,138^, Lili Milani^137^, Vihra Milanova^139^, Younes Mokrab^8^, Derek W. Morris^5,63^, Ole Mors^17,58,140^, Kieran C. Murphy^141^, Robin M. Murray^142^, Inez Myin-Germeys^143^, Bertram Müller-Myhsok^144,145,146^, Mari Nelis^137^, Igor Nenadic^147^, Deborah A. Nertney^148^, Gerald Nestadt^149^, Kristin K. Nicodemus^150^, Liene Nikitina-Zake^110^, Laura Nisenbaum^151^, Annelie Nordin^152^, Eadbhard O’Callaghan^153^, Colm O’Dushlaine^2^, F. Anthony O’Neill^154^, Sang-Yun Oh^155^, Ann Olincy^79^, Line Olsen^17,90^, Jim Van Os^143,156^, Psychosis Endophenotypes International Consortium^157^, Christos Pantelis^39,158^, George N. Papadimitriou^60^, Sergi Papiol^23^, Elena Parkhomenko^36^, Michele T. Pato^111^, Tiina Paunio^159,160^, Milica Pejovic-Milovancevic^161^, Diana O. Perkins^162^, Olli Pietiläinen^160,163^, Jonathan Pimm^53^, Andrew J. Pocklington^6^, John Powell^142^, Alkes Price^164^, Ann E. Pulver^149^, Shaun M. Purcell^82^, Digby Quested^165^, Henrik B. Rasmussen^17,90^, Abraham Reichenberg^36^, Mark A. Reimers^166^, Alexander L. Richards^6,7^, Joshua L. Roffman^30,32^, Panos Roussos^82,167^, Douglas M. Ruderfer^82^, Veikko Salomaa^71^, Alan R. Sanders^64,65^, Ulrich Schall^39,122^, Christian R. Schubert^168^, Thomas G. Schulze^77,169^, Sibylle G. Schwab^170^, Edward M. Scolnick^2^, Rodney J. Scott^39,171,172^, Larry J. Seidman^130,136^, Jianxin Shi^173^, Engilbert Sigurdsson^174^, Teimuraz Silagadze^175^, Jeremy M. Silverman^36,176^, Kang Sim^47^, Petr Slominsky^109^, Jordan W. Smoller^2,4^, Hon-Cheong So^43^, Chris C. A. Spencer^177^, Eli A. Stahl^3,82^, Hreinn Stefansson^178^, Stacy Steinberg^178^, Elisabeth Stogmann^179^, Richard E. Straub^180^, Eric Strengman^181,182^, Jana Strohmaier^77^, T. Scott Stroup^121^, Mythily Subramaniam^47^, Jaana Suvisaari^124^, Dragan M. Svrakic^48^, Jin P. Szatkiewicz^51^, Erik Söderman^12^, Srinivas Thirumalai^183^, Draga Toncheva^104^, Sarah Tosato^184^, Juha Veijola^185,186^, John Waddington^187^, Dermot Walsh^188^, Dai Wang^86^, Qiang Wang^119^, Bradley T. Webb^22^, Mark Weiser^54^, Dieter B. Wildenauer^189^, Nigel M. Williams^190^, Stephanie Williams^51^, Stephanie H. Witt^77^, Aaron R. Wolen^166^, Emily H. M. Wong^43^, Brandon K. Wormley^22^, Hualin Simon Xi^191^, Clement C. Zai^106,107^, Xuebin Zheng^192^, Fritz Zimprich^179^, Naomi R. Wray^87^, Kari Stefansson^178^, Peter M. Visscher^87^, Wellcome Trust Case-Control Consortium 2^193^, Rolf Adolfsson^152^, Ole A. Andreassen^14,135^, Douglas H. R. Blackwood^134^, Elvira Bramon^194^, Joseph D. Buxbaum^35,36,91,195^, Anders D. Børglum^17,58,59,140^, Sven Cichon^55,56,95,196^, Ariel Darvasi^197^, Enrico Domenici^198^, Hannelore Ehrenreich^23^, Tõnu Esko^3,11,96,137^, Pablo V. Gejman^64,65^, Michael Gill^5^, Hugh Gurling^53^, Christina M. Hultman^26^, Nakao Iwata^98^, Assen V. Jablensky^39,199,200,201^, Erik G. Jönsson^12^, Kenneth S. Kendler^202^, George Kirov^6^, Jo Knight^106,107,108^, Todd Lencz^203,204,205^, Douglas F. Levinson^19^, Qingqin S. Li^86^, Jianjun Liu^192,206^, Anil K. Malhotra^203,204,205^, Steven A. McCarroll^2,96^, Andrew McQuillin^53^, Jennifer L. Moran^2^, Preben B. Mortensen^15,16,17^, Bryan J. Mowry^87,207^, Markus M. Nöthen^55,56^, Roel A. Ophoff^38,80,208^, Michael J. Owen^6,7^, Aarno Palotie^4,163,209^, Carlos N. Pato^111^, Tracey L. Petryshen^130,209,210^, Danielle Posthuma^211,212,213^, Marcella Rietschel^77^, Brien P. Riley^202^, Dan Rujescu^81,83^, Pak C. Sham^214^, Pamela Sklar^82,91,167^, David St Clair^215^, Daniel R. Weinberger^180,216^, Jens R. Wendland^168^, Thomas Werge^17,90,217^, Mark J. Daly^1^, Patrick F. Sullivan^26,51,162^ & Michael C. O’Donovan^6,7^

^1^Analytic and Translational Genetics Unit, Massachusetts General Hospital, Boston, Massachusetts, USA.

^2^Stanley Center for Psychiatric Research, Broad Institute of MIT and Harvard, Cambridge, Massachusetts, USA.

^3^Medical and Population Genetics Program, Broad Institute of MIT and Harvard, Cambridge, Massachusetts, USA.

^4^Psychiatric and Neurodevelopmental Genetics Unit, Massachusetts General Hospital, Boston, Massachusetts, USA.

^5^Neuropsychiatric Genetics Research Group, Department of Psychiatry, Trinity College Dublin, Ireland.

^6^MRC Centre for Neuropsychiatric Genetics and Genomics, Institute of Psychological Medicine and Clinical Neurosciences, School of Medicine, Cardiff University, Cardiff, UK. ^7^National Centre for Mental Health, Cardiff University, Cardiff, Wales.

^8^Eli Lilly and Company Limited, Erl Wood Manor, Sunninghill Road, Windlesham, Surrey, UK.

^9^Social, Genetic and Developmental Psychiatry Centre, Institute of Psychiatry, King’s College London, London, UK.

^10^Center for Biological Sequence Analysis, Department of Systems Biology, Technical University of Denmark, Lyngby, Denmark.

^11^Division of Endocrinology and Center for Basic and Translational Obesity Research, Boston Children’s Hospital, Boston, Massachusetts, USA.

^12^Department of Clinical Neuroscience, Karolinska Institutet, Stockholm, Sweden.

^13^Department of Psychiatry, Diakonhjemmet Hospital, Oslo, Norway.

^14^NORMENT, KG Jebsen Centre for Psychosis Research, Institute of Clinical Medicine, University of Oslo, Oslo, Norway.

^15^Centre for Integrative Register-based Research, CIRRAU, Aarhus University, Aarhus, Denmark.

^16^National Centre for Register-based Research, Aarhus University, Aarhus, Denmark. ^17^The Lundbeck Foundation Initiative for Integrative Psychiatric Research, iPSYCH, Denmark.

^18^State Mental Hospital, Haar, Germany.

^19^Department of Psychiatry and Behavioral Sciences, Stanford University, Stanford, California, USA.

^20^Department of Psychiatry and Behavioral Sciences, Atlanta Veterans Affairs Medical Center, Atlanta, Georgia, USA.

^21^Department of Psychiatry and Behavioral Sciences, Emory University, Atlanta, Georgia, USA.

^22^Virginia Institute for Psychiatric and Behavioral Genetics, Department of Psychiatry, Virginia Commonwealth University, Richmond, Virginia, USA.

^23^Clinical Neuroscience, Max Planck Institute of Experimental Medicine, Göttingen, Germany.

^24^Department of Medical Genetics, University of Pécs, Pécs, Hungary.

^25^Szentagothai Research Center, University of Pécs, Pécs, Hungary.

^26^Department of Medical Epidemiology and Biostatistics, Karolinska Institutet, Stockholm, Sweden.

^27^Department of Psychiatry, University of Iowa Carver College of Medicine, Iowa City, Iowa, USA.

^28^University Medical Center Groningen, Department of Psychiatry, University of Groningen, The Netherlands.

^29^School of Nursing, Louisiana State University Health Sciences Center, New Orleans, Louisiana, USA.

^30^Athinoula A. Martinos Center, Massachusetts General Hospital, Boston, Massachusetts, USA.

^31^Center for Brain Science, Harvard University, Cambridge, Massachusetts, USA. ^32^Department of Psychiatry, Massachusetts General Hospital, Boston, Massachusetts, USA. ^33^Department of Psychiatry, University of California at San Francisco, San Francisco, California, USA.

^34^University Medical Center Utrecht, Department of Psychiatry, Rudolf Magnus Institute of Neuroscience, The Netherlands.

^35^Department of Human Genetics, Icahn School of Medicine at Mount Sinai, New York, New York, USA.

^36^Department of Psychiatry, Icahn School of Medicine at Mount Sinai, New York, New York, USA.

^37^Centre Hospitalier du Rouvray and INSERM U1079 Faculty of Medicine, Rouen, France. ^38^Department of Human Genetics, David Geffen School of Medicine, University of California, Los Angeles, California, USA.

^39^Schizophrenia Research Institute, Sydney, Australia.

^40^School of Psychiatry, University of New South Wales, Sydney, Australia.

^41^Royal Brisbane and Women’s Hospital, University of Queensland, Brisbane, Australia.

^42^Institute of Psychology, Chinese Academy of Science, Beijing, China.

^43^Department of Psychiatry, Li Ka Shing Faculty of Medicine, The University of Hong Kong, Hong Kong, China.

^44^Department of Psychiatry and State Ket Laboratory for Brain and Cognitive Sciences, Li Ka Shing Faculty of Medicine, The University of Hong Kong, Hong Kong, China.

^45^Department of Computer Science, University of North Carolina, Chapel Hill, North Carolina, USA.

^46^Castle Peak Hospital, Hong Kong, China.

^47^Institute of Mental Health, Singapore.

^48^Department of Psychiatry, Washington University, St. Louis, Missouri, USA. ^49^Department of Child and Adolescent Psychiatry, Pierre and Marie Curie Faculty of Medicine and Brain and Spinal Cord Institute (ICM), Paris, France.

^50^Neuroscience Therapeutic Area, Janssen Research and Development, Raritan, New Jersey, USA

^51^Department of Genetics, University of North Carolina, Chapel Hill, North Carolina, USA.

^52^Department of Psychological Medicine, Queen Mary University of London, UK. ^53^Molecular Psychiatry Laboratory, Division of Psychiatry, University College London, UK. ^54^Sheba Medical Center, Tel Hashomer, Israel.

^55^Department of Genomics, Life and Brain Center, Bonn, Germany.

^56^Institute of Human Genetics, University of Bonn, Bonn, Germany.

^57^Applied Molecular Genomics Unit, VIB Department of Molecular Genetics, University of Antwerp, Antwerp, Belgium.

^58^Centre for Integrative Sequencing, iSEQ, Aarhus University, Aarhus, Denmark.

^59^Department of Biomedicine, Aarhus University, Aarhus, Denmark.

^60^First Department of Psychiatry, University of Athens Medical School, Athens, Greece.

^61^Department of Psychiatry, University College Cork, Ireland.

^62^Department of Medical Genetics, Oslo University Hospital, Oslo, Norway. ^63^Cognitive Genetics and Therapy Group, School of Psychology and Discipline of Biochemistry, National University of Ireland Galway, Ireland.

^64^Department of Psychiatry and Behavioral Neuroscience, University of Chicago, Chicago, Illinois, USA.

^65^Department of Psychiatry and Behavioral Sciences, NorthShore University HealthSystem, Evanston, Illinois, USA.

^66^Department of Non-Communicable Disease Epidemiology, London School of Hygiene and Tropical Medicine, London, UK.

^67^Department of Child and Adolescent Psychiatry, University Clinic of Psychiatry, Skopje, Republic of Macedonia. ^68^Department of Psychiatry, University of Regensburg, Regensburg, Germany.

^69^Department of General Practice, Helsinki University Central Hospital, Helsinki, Finland.

^70^Folkhälsan Research Center, Helsinki, Finland.

^71^National Institute for Health and Welfare, Helsinki, Finland.

^72^Translational Technologies and Bioinformatics, Pharma Research and Early Development, F. Hoffman-La Roche, Basel, Switzerland.

^73^Department of Psychiatry, Georgetown University School of Medicine, Washington DC, USA.

^74^Department of Psychiatry, Keck School of Medicine of the University of Southern California, Los Angeles, California, USA.

^75^Department of Psychiatry, Virginia Commonwealth University School of Medicine, Richmond, Virginia, USA.

^76^Mental Health Service Line, Washington VA Medical Center, Washington DC, USA. ^77^Department of Genetic Epidemiology in Psychiatry, Central Institute of Mental Health, Medical Faculty Mannheim, University of Heidelberg, Heidelberg, Germany.

^78^Department of Genetics, University of Groningen, University Medical Centre Groningen, The Netherlands.

^79^Department of Psychiatry, University of Colorado Denver, Aurora, Colorado, USA. ^80^Center for Neurobehavioral Genetics, Semel Institute for Neuroscience and Human Behavior, University of California, Los Angeles, California, USA.

^81^Department of Psychiatry, University of Halle, Halle, Germany.

^82^Division of Psychiatric Genomics, Department of Psychiatry, Icahn School of Medicine at Mount Sinai, New York, New York, USA.

^83^Department of Psychiatry, University of Munich, Munich, Germany.

^84^Departments of Psychiatry and Human and Molecular Genetics, INSERM, Institut de Myologie, Hôpital de la Pitiè-Salpêtrière, Paris, France.

^85^Mental Health Research Centre, Russian Academy of Medical Sciences, Moscow, Russia. ^86^Neuroscience Therapeutic Area, Janssen Research and Development, Raritan, New Jersey, USA.

^87^Queensland Brain Institute, The University of Queensland, Brisbane, Queensland, Australia.

^88^Academic Medical Centre University of Amsterdam, Department of Psychiatry, Amsterdam, The Netherlands.

^89^Illumina, La Jolla, California, USA.

^90^Institute of Biological Psychiatry, MHC Sct. Hans, Mental Health Services Copenhagen, Denmark.

^91^Friedman Brain Institute, Icahn School of Medicine at Mount Sinai, New York, New York, USA.

^92^J. J. Peters VA Medical Center, Bronx, New York, USA.

^93^Priority Research Centre for Health Behaviour, University of Newcastle, Newcastle, Australia.

^94^School of Electrical Engineering and Computer Science, University of Newcastle, Newcastle, Australia.

^95^Division of Medical Genetics, Department of Biomedicine, University of Basel, Basel, Switzerland.

^96^Department of Genetics, Harvard Medical School, Boston, Massachusetts, USA. ^97^Section of Neonatal Screening and Hormones, Department of Clinical Biochemistry, Immunology and Genetics, Statens Serum Institut, Copenhagen, Denmark.

^98^Department of Psychiatry, Fujita Health University School of Medicine, Toyoake, Aichi, Japan.

^99^Regional Centre for Clinical Research in Psychosis, Department of Psychiatry, Stavanger University Hospital, Stavanger, Norway.

^100^Rheumatology Research Group, Vall d’Hebron Research Institute, Barcelona, Spain. ^101^Department of Psychiatry, Rudolf Magnus Institute of Neuroscience, University Medical Center Utrecht, Utrecht, The Netherlands.

^102^Centre for Medical Research, The University of Western Australia, Perth, Western Australia, Australia.

^103^Perkins Institute for Medical Research, The University of Western Australia, Perth, Western Australia, Australia.

^104^Department of Medical Genetics, Medical University, Sofia, Bulgaria.

^105^Department of Psychology, University of Colorado Boulder, Boulder, Colorado, USA. ^106^Campbell Family Mental Health Research Institute, Centre for Addiction and Mental Health, Toronto, Ontario, Canada.

^107^Department of Psychiatry, University of Toronto, Toronto, Ontario, Canada.

^108^Institute of Medical Science, University of Toronto, Toronto, Ontario, Canada.

^109^Institute of Molecular Genetics, Russian Academy of Sciences, Moscow, Russia.

^110^Latvian Biomedical Research and Study Centre, Riga, Latvia.

^111^Department of Psychiatry and Zilkha Neurogenetics Institute, Keck School of Medicine at University of Southern California, Los Angeles, California, USA.

^112^Faculty of Medicine, Vilnius University, Vilnius, Lithuania.

^113^2nd Faculty of Medicine and University Hospital Motol, Prague, Czech Republic. ^114^Department of Biology and Medical Genetics, Charles University Prague, Prague, Czech Republic.

^115^Pierre and Marie Curie Faculty of Medicine, Paris, France.

^116^Duke-NUS Graduate Medical School, Singapore.

^117^Department of Psychiatry, Hadassah-Hebrew University Medical Center, Jerusalem, Israel. ^118^Centre for Genomic Sciences and Department of Psychiatry, Li Ka Shing Faculty of Medicine, The University of Hong Kong, Hong Kong, China.

^119^Mental Health Centre and Psychiatric Laboratory, West China Hospital, Sichuan University, Chendu, Sichuan, China.

^120^Department of Biostatistics, Johns Hopkins University Bloomberg School of Public Health, Baltimore, Maryland, USA.

^121^Department of Psychiatry, Columbia University, New York, New York, USA. ^122^Priority Centre for Translational Neuroscience and Mental Health, University of Newcastle, Newcastle, Australia.

^123^Department of Genetics and Pathology, International Hereditary Cancer Center, Pomeranian Medical University in Szczecin, Szczecin, Poland.

^124^Department of Mental Health and Substance Abuse Services; National Institute for Health and Welfare, Helsinki, Finland.

^125^Department of Mental Health, Bloomberg School of Public Health, Johns Hopkins University, Baltimore, Maryland, USA.

^126^Department of Psychiatry, University of Bonn, Bonn, Germany.

^127^Centre National de la Recherche Scientifique, Laboratoire de Génétique Moléculaire de la Neurotransmission et des Processus Neurodégénératifs, Hôpital de la Pitié Salpêtrière, Paris, France.

^128^Department of Genomics Mathematics, University of Bonn, Bonn, Germany.

^129^Research Unit, Sørlandet Hospital, Kristiansand, Norway.

^130^Department of Psychiatry, Harvard Medical School, Boston, Massachusetts, USA.

^131^VA Boston Health Care System, Brockton, Massachusetts, USA.

^132^Department of Psychiatry, National University of Ireland Galway, Ireland.

^133^Centre for Cognitive Ageing and Cognitive Epidemiology, University of Edinburgh, UK.

^134^Division of Psychiatry, University of Edinburgh, Edinburgh, UK.

^135^Division of Mental Health and Addiction, Oslo University Hospital, Oslo, Norway. ^136^Massachusetts Mental Health Center Public Psychiatry Division of the Beth Israel Deaconess Medical Center, Boston, Massachusetts, USA.

^137^Estonian Genome Center, University of Tartu, Tartu, Estonia. ^138^School of Psychology, University of Newcastle, Newcastle, Australia. ^139^First Psychiatric Clinic, Medical University, Sofia, Bulgaria.

^140^Department P, Aarhus University Hospital, Risskov, Denmark. ^141^Department of Psychiatry, Royal College of Surgeons in Ireland, Ireland. ^142^King’s College London, UK.

^143^Maastricht University Medical Centre, South Limburg Mental Health Research and Teaching Network, EURON, Maastricht, The Netherlands.

^144^Institute of Translational Medicine, University Liverpool, UK.

^145^Max Planck Institute of Psychiatry, Munich, Germany.

^146^Munich Cluster for Systems Neurology (SyNergy), Munich, Germany.

^147^Department of Psychiatry and Psychotherapy, Jena University Hospital, Jena, Germany. ^148^Department of Psychiatry, Queensland Brain Institute and Queensland Centre for Mental Health Research, University of Queensland, Brisbane, Queensland, Australia.

^149^Department of Psychiatry and Behavioral Sciences, Johns Hopkins University School of Medicine, Baltimore, Maryland, USA.

^150^Department of Psychiatry, Trinity College Dublin, Ireland.

^151^Eli Lilly and Company, Lilly Corporate Center, Indianapolis, Indiana, USA. ^152^Department of Clinical Sciences, Psychiatry, Umeå University, Umeå, Sweden. ^153^DETECT Early Intervention Service for Psychosis, Blackrock, Dublin, Ireland. ^154^Centre for Public Health, Institute of Clinical Sciences, Queen’s University Belfast, Belfast, UK.

^155^Lawrence Berkeley National Laboratory, University of California at Berkeley, Berkeley, California, USA.

^156^Institute of Psychiatry at King’s College London, London, UK.

^157^A list of authors and affiliations appears in the Supplementary Information. ^158^Melbourne Neuropsychiatry Centre, University of Melbourne & Melbourne Health, Melbourne, Australia.

^159^Department of Psychiatry, University of Helsinki, Finland.

^160^Public Health Genomics Unit, National Institute for Health and Welfare, Helsinki, Finland.

^161^Medical Faculty, University of Belgrade, Belgrade, Serbia.

^162^Department of Psychiatry, University of North Carolina, Chapel Hill, North Carolina, USA.

^163^Institute for Molecular Medicine Finland, FIMM, Helsinki, Finland. ^164^Department of Epidemiology, Harvard University, Boston, Massachusetts, USA. ^165^Department of Psychiatry, University of Oxford, Oxford, UK.

^166^Virginia Institute for Psychiatric and Behavioral Genetics, Virginia Commonwealth University, Richmond, Virginia, USA.

^167^Institute for Multiscale Biology, Icahn School of Medicine at Mount Sinai, New York, New York, USA.

^168^PharmaTherapeutics Clinical Research, Pfizer Worldwide Research and Development, Cambridge, Massachusetts, USA.

^169^Department of Psychiatry and Psychotherapy, University of Gottingen, Göttingen, Germany.

^170^Psychiatry and Psychotherapy Clinic, University of Erlangen, Germany.

^171^Hunter New England Health Service, Newcastle, Australia.

^172^School of Biomedical Sciences, University of Newcastle, Newcastle, Australia. ^173^Division of Cancer Epidemiology and Genetics, National Cancer Institute, Bethesda, Maryland, USA.

^174^University of Iceland, Landspitali, National University Hospital, Reykjavik, Iceland. ^175^Department of Psychiatry and Drug Addiction, Tbilisi State Medical University (TSMU), Tbilisi, Georgia.

^176^Research and Development, Bronx Veterans Affairs Medical Center, New York, New York, USA.

^177^Wellcome Trust Centre for Human Genetics, Oxford, UK.

^178^deCODE Genetics, Reykjavik, Iceland.

^179^Department of Clinical Neurology, Medical University of Vienna, Austria.

^180^Lieber Institute for Brain Development, Baltimore, Maryland, USA.

^181^Department of Medical Genetics, University Medical Centre, Utrecht, The Netherlands.

^182^Rudolf Magnus Institute of Neuroscience, University Medical Centre Utrecht, The Netherlands.

^183^Berkshire Healthcare NHS Foundation Trust, Bracknell, UK.

^184^Section of Psychiatry, University of Verona, Verona, Italy.

^185^Department of Psychiatry, University of Oulu, Finland.

^186^University Hospital of Oulu, Oulu, Finland.

^187^Molecular and Cellular Therapeutics, Royal College of Surgeons in Ireland, Dublin, Ireland.

^188^Health Research Board, Dublin, Ireland.

^189^Department of Psychiatry and Clinical Neurosciences, School of Psychiatry and Clinical Neurosciences, Queen Elizabeth II Medical Centre, Perth, Western Australia, Australia.

^190^Department of Psychological Medicine and Neurology, MRC Centre for Neuropsychiatric Genetics and Genomics, School of Medicine, Cardiff University, Cardiff, Wales, UK. ^191^Computational Sciences CoE, Pfizer Worldwide Research and Development, Cambridge, Massachusetts, USA.

^192^Human Genetics, Genome Institute of Singapore, A*STAR, Singapore. ^193^A list of authors and affiliations appears in the Supplementary Information. ^194^University College London, UK.

^195^Department of Neuroscience, Icahn School of Medicine at Mount Sinai, New York, New York, USA.

^196^Institute of Neuroscience and Medicine (INM-1), Research Center Juelich, Juelich, Germany.

^197^Department of Genetics, The Hebrew University of Jerusalem, Jerusalem, Israel. ^198^Neuroscience Discovery and Translational Area, Pharma Research and Early Development, F. Hoffman-La Roche, Basel, Switzerland.

^199^School of Psychiatry and Clinical Neurosciences, The University of Western Australia, Perth, Australia.

^200^The Perkins Institute of Medical Research, Perth, Australia.

^201^UWA Centre for Clinical Research in Neuropsychiatry, Crawley 6009, Western Australia. ^202^Virginia Institute for Psychiatric and Behavioral Genetics 23298-980126, Departments of Psychiatry and Human and Molecular Genetics 23298-980003, Virginia Commonwealth University, Richmond, Virginia, USA.

^203^The Feinstein Institute for Medical Research, Manhasset, New York, 11030 USA.

^204^The Hofstra NS-LIJ School of Medicine, Hempstead, New York, 11549 USA.

^205^The Zucker Hillside Hospital, Glen Oaks, New York,11004 USA.

^206^Saw Swee Hock School of Public Health, National University of Singapore, 117597 Singapore.

^207^Queensland Centre for Mental Health Research, University of Queensland, Brisbane 4076, Queensland, Australia.

^208^Department of Psychiatry, Brain Center Rudolf Magnus, University Medical Center Utrecht,3584 The Netherlands.

^209^The Broad Institute of MIT and Harvard, Cambridge, Massachusetts 02142, USA. ^210^Center for Human Genetic Research and Department of Psychiatry, Massachusetts General Hospital, Boston, Massachusetts 02114, USA.

^211^Department of Child and Adolescent Psychiatry, Erasmus University Medical Centre, Rotterdam 3000, The Netherlands.

^212^Department of Complex Trait Genetics, Neuroscience Campus Amsterdam, VU University Medical Center Amsterdam, Amsterdam 1081, The Netherlands.

^213^Department of Functional Genomics, Center for Neurogenomics and Cognitive Research, Neuroscience Campus Amsterdam, VU University, Amsterdam 1081, The Netherlands.

^214^Centre for Genomic Sciences, State Ket Laboratory for Brain and Cognitive Sciences, and Department of Psychiatry, Li Ka Shing Faculty of Medicine, The University of Hong Kong, Hong Kong, China. n/a

^215^University of Aberdeen, Institute of Medical Sciences, Aberdeen, Scotland AB25 2ZD, UK.

^216^Departments of Psychiatry, Neurology, Neuroscience and Institute of Genetic Medicine, Johns Hopkins School of Medicine, Baltimore, Maryland 21287-7413, USA.

^217^Department of Clinical Medicine, University of Copenhagen, Copenhagen 2200, Denmark.

### Major Depressive Disorder Working Group of the Psychiatric Genomics Consortium

Naomi R Wray* ^1, 2^, Stephan Ripke* ^3, 4, 5^, Manuel Mattheisen* ^6, 7, 8^, Maciej Trzaskowski ^1^, Enda M Byrne ^1^, Abdel Abdellaoui ^9^, Mark J Adams ^10^, Esben Agerbo ^11, 12, 13^, Tracy M Air ^14^, Till F M Andlauer ^15, 16^, Silviu-Alin Bacanu ^17^, Marie Bækvad-Hansen ^13, 18^, Aartjan T F Beekman ^19^, Tim B Bigdeli ^17, 20^, Elisabeth B Binder ^15, 21^, Julien Bryois ^22^, Henriette N Buttenschøn ^13, 23, 24^, Jonas Bybjerg-Grauholm ^13, 18^, Na Cai ^25, 26^, Enrique Castelao ^27^, Jane Hvarregaard Christensen ^8, 13, 24^, Toni-Kim Clarke ^10^, Jonathan R I Coleman ^28^, Lucía Colodro-Conde ^29^, Baptiste Couvy-Duchesne ^2, 30^, Nick Craddock ^31^, Gregory E Crawford ^32, 33^, Gail Davies ^34^, Ian J Deary ^34^, Franziska Degenhardt ^35^, Eske M Derks ^29^, Nese Direk ^36, 37^, Conor V Dolan ^9^, Erin C Dunn ^38, 39, 40^, Thalia C Eley ^28^, Valentina Escott-Price ^41^, Farnush Farhadi Hassan Kiadeh ^42^, Hilary K Finucane ^43, 44^, Jerome C Foo ^45^, Andreas J Forstner ^35, 46, 47, 48^, Josef Frank ^45^, Héléna A Gaspar ^28^, Michael Gill ^49^, Fernando S Goes ^50^, Scott D Gordon ^29^, Jakob Grove ^8, 13, 24, 51^, Lynsey S Hall ^10, 52^, Christine Søholm Hansen ^13, 18^, Thomas F Hansen ^53, 54, 55^, Stefan Herms ^35, 47^, Ian B Hickie ^56^, Per Hoffmann ^35, 47^, Georg Homuth ^57^, Carsten Horn ^58^, Jouke-Jan Hottenga ^9^, David M Hougaard ^13, 18^, David M Howard ^10, 28^, Marcus Ising ^59^, Rick Jansen ^19^, Ian Jones ^60^, Lisa A Jones ^61^, Eric Jorgenson ^62^, James A Knowles ^63^, Isaac S Kohane ^64, 65, 66^, Julia Kraft ^4^, Warren W. Kretzschmar ^67^, Zoltán Kutalik ^68, 69^, Yihan Li ^67^, Penelope A Lind ^29^, Donald J MacIntyre ^70, 71^, Dean F MacKinnon ^50^, Robert M Maier ^2^, Wolfgang Maier ^72^, Jonathan Marchini ^73^, Hamdi Mbarek ^9^, Patrick McGrath ^74^, Peter McGuffin ^28^, Sarah E Medland ^29^, Divya Mehta ^2, 75^, Christel M Middeldorp ^9, 76, 77^, Evelin Mihailov ^78^, Yuri Milaneschi ^19^, Lili Milani ^78^, Francis M Mondimore ^50^, Grant W Montgomery ^1^, Sara Mostafavi ^79, 80^, Niamh Mullins ^28^, Matthias Nauck ^81, 82^, Bernard Ng ^80^, Michel G Nivard ^9^, Dale R Nyholt ^83^, Paul F O’Reilly ^28^, Hogni Oskarsson ^84^, Michael J Owen ^60^, Jodie N Painter ^29^, Carsten Bøcker Pedersen ^11, 12, 13^, Marianne Giørtz Pedersen ^11, 12, 13^, Roseann E Peterson ^17, 85^, Wouter J Peyrot ^19^, Giorgio Pistis ^27^, Danielle Posthuma ^86, 87^, Jorge A Quiroz ^88^, Per Qvist ^8, 13, 24^, John P Rice ^89^, Brien P. Riley ^17^, Margarita Rivera ^28, 90^, Saira Saeed Mirza ^36^, Robert Schoevers ^91^, Eva C Schulte ^92, 93^, Ling Shen ^62^, Jianxin Shi ^94^, Stanley I Shyn ^95^, Engilbert Sigurdsson ^96^, Grant C B Sinnamon ^97^, Johannes H Smit ^19^, Daniel J Smith ^98^, Hreinn Stefansson ^99^, Stacy Steinberg ^99^, Fabian Streit ^45^, Jana Strohmaier ^45^, Katherine E Tansey ^100^, Henning Teismann ^101^, Alexander Teumer ^102^, Wesley Thompson ^13, 54, 103, 104^, Pippa A Thomson ^105^, Thorgeir E Thorgeirsson ^99^, Matthew Traylor ^106^, Jens Treutlein ^45^, Vassily Trubetskoy ^4^, André G Uitterlinden ^107^, Daniel Umbricht ^108^, Sandra Van der Auwera ^109^, Albert M van Hemert ^110^, Alexander Viktorin ^22^, Peter M Visscher ^1, 2^, Yunpeng Wang ^13, 54, 104^, Bradley T. Webb ^111^, Shantel Marie Weinsheimer ^13, 54^, Jürgen Wellmann ^101^, Gonneke Willemsen ^9^, Stephanie H Witt ^45^, Yang Wu ^1^, Hualin S Xi ^112^, Jian Yang ^2, 113^, Futao Zhang ^1^, Volker Arolt ^114^, Bernhard T Baune ^114, 115, 116^, Klaus Berger ^101^, Dorret I Boomsma ^9^, Sven Cichon ^35, 47, 117, 118^, Udo Dannlowski ^114^, EJC de Geus ^9, 119^, J Raymond DePaulo ^50^, Enrico Domenici ^120^, Katharina Domschke ^121, 122^, Tõnu Esko ^5, 78^, Hans J Grabe ^109^, Steven P Hamilton ^123^, Caroline Hayward ^124^, Andrew C Heath ^89^, Kenneth S Kendler ^17^, Stefan Kloiber ^59, 125, 126^, Glyn Lewis ^127^, Qingqin S Li ^128^, Susanne Lucae ^59^, Pamela AF Madden ^89^, Patrik K Magnusson ^22^, Nicholas G Martin ^29^, Andrew M McIntosh ^10, 34^, Andres Metspalu ^78, 129^, Ole Mors ^13, 130^, Preben Bo Mortensen ^11, 12, 13, 24^, Bertram Müller-Myhsok ^15, 131, 132^, Merete Nordentoft ^13, 133^, Markus M Nöthen ^35^, Michael C O’Donovan ^60^, Sara A Paciga ^134^, Nancy L Pedersen ^22^, Brenda WJH Penninx ^19^, Roy H Perlis ^38, 135^, David J Porteous ^105^, James B Potash ^136^, Martin Preisig ^27^, Marcella Rietschel ^45^, Catherine Schaefer ^62^, Thomas G Schulze ^45, 93, 137, 138, 139^, Jordan W Smoller ^38, 39, 40^, Kari Stefansson ^99, 140^, Henning Tiemeier ^36, 141, 142^, Rudolf Uher ^143^, Henry Völzke ^102^, Myrna M Weissman ^74, 144^, Thomas Werge ^13, 54, 145^, Cathryn M Lewis* ^28, 146^, Douglas F Levinson* ^147^, Gerome Breen* ^28, 148^, Anders D Børglum* ^8, 13, 24^, Patrick F Sullivan* ^22, 149, 150^

1, Institute for Molecular Bioscience, The University of Queensland, Brisbane, QLD, AU 2, Queensland Brain Institute, The University of Queensland, Brisbane, QLD, AU

3, Analytic and Translational Genetics Unit, Massachusetts General Hospital, Boston, MA, US

4, Department of Psychiatry and Psychotherapy, Universitätsmedizin Berlin Campus Charité Mitte, Berlin, DE

5, Medical and Population Genetics, Broad Institute, Cambridge, MA, US

6, Department of Psychiatry, Psychosomatics and Psychotherapy, University of Wurzburg, Wurzburg, DE

7, Centre for Psychiatry Research, Department of Clinical Neuroscience, Karolinska Institutet, Stockholm, SE

8, Department of Biomedicine, Aarhus University, Aarhus, DK

9, Dept of Biological Psychology & EMGO+ Institute for Health and Care Research, Vrije Universiteit Amsterdam, Amsterdam, NL

10, Division of Psychiatry, University of Edinburgh, Edinburgh, GB

11, Centre for Integrated Register-based Research, Aarhus University, Aarhus, DK 12, National Centre for Register-Based Research, Aarhus University, Aarhus, DK

13, iPSYCH, The Lundbeck Foundation Initiative for Integrative Psychiatric Research,, DK 14, Discipline of Psychiatry, University of Adelaide, Adelaide, SA, AU

15, Department of Translational Research in Psychiatry, Max Planck Institute of Psychiatry, Munich, DE

16, Department of Neurology, Klinikum rechts der Isar, Technical University of Munich, Munich, DE

17, Department of Psychiatry, Virginia Commonwealth University, Richmond, VA, US 18, Center for Neonatal Screening, Department for Congenital Disorders, Statens Serum Institut, Copenhagen, DK

19, Department of Psychiatry, Vrije Universiteit Medical Center and GGZ inGeest, Amsterdam, NL

20, Virginia Institute for Psychiatric and Behavior Genetics, Richmond, VA, US 21, Department of Psychiatry and Behavioral Sciences, Emory University School of Medicine, Atlanta, GA, US

22, Department of Medical Epidemiology and Biostatistics, Karolinska Institutet, Stockholm, SE

23, Department of Clinical Medicine, Translational Neuropsychiatry Unit, Aarhus University, Aarhus, DK

24, iSEQ, Centre for Integrative Sequencing, Aarhus University, Aarhus, DK 25, Human Genetics, Wellcome Trust Sanger Institute, Cambridge, GB

26, Statistical genomics and systems genetics, European Bioinformatics Institute (EMBL-EBI), Cambridge, GB

27, Department of Psychiatry, Lausanne University Hospital and University of Lausanne, Lausanne, CH

28, Social, Genetic and Developmental Psychiatry Centre, King’s College London, London, GB

29, Genetics and Computational Biology, QIMR Berghofer Medical Research Institute, Brisbane, QLD, AU

30, Centre for Advanced Imaging, The University of Queensland, Brisbane, QLD, AU 31, Psychological Medicine, Cardiff University, Cardiff, GB

32, Center for Genomic and Computational Biology, Duke University, Durham, NC, US 33, Department of Pediatrics, Division of Medical Genetics, Duke University, Durham, NC, US

34, Centre for Cognitive Ageing and Cognitive Epidemiology, University of Edinburgh, Edinburgh, GB

35, Institute of Human Genetics, University of Bonn, School of Medicine & University Hospital Bonn, Bonn, DE

36, Epidemiology, Erasmus MC, Rotterdam, Zuid-Holland, NL

37, Psychiatry, Dokuz Eylul University School Of Medicine, Izmir, TR

38, Department of Psychiatry, Massachusetts General Hospital, Boston, MA, US

39, Psychiatric and Neurodevelopmental Genetics Unit (PNGU), Massachusetts General Hospital, Boston, MA, US

40, Stanley Center for Psychiatric Research, Broad Institute, Cambridge, MA, US 41, Neuroscience and Mental Health, Cardiff University, Cardiff, GB

42, Bioinformatics, University of British Columbia, Vancouver, BC, CA

43, Department of Epidemiology, Harvard T.H. Chan School of Public Health, Boston, MA, US

44, Department of Mathematics, Massachusetts Institute of Technology, Cambridge, MA, US 45, Department of Genetic Epidemiology in Psychiatry, Central Institute of Mental Health, Medical Faculty Mannheim, Heidelberg University, Mannheim, Baden-Württemberg, DE

46, Department of Psychiatry (UPK), University of Basel, Basel, CH 47, Department of Biomedicine, University of Basel, Basel, CH

48, Centre for Human Genetics, University of Marburg, Marburg, DE 49, Department of Psychiatry, Trinity College Dublin, Dublin, IE

50, Psychiatry & Behavioral Sciences, Johns Hopkins University, Baltimore, MD, US 51, Bioinformatics Research Centre, Aarhus University, Aarhus, DK

52, Institute of Genetic Medicine, Newcastle University, Newcastle upon Tyne, GB 53, Danish Headache Centre, Department of Neurology, Rigshospitalet, Glostrup, DK 54, Institute of Biological Psychiatry, Mental Health Center Sct. Hans, Mental Health Services Capital Region of Denmark, Copenhagen, DK

55, iPSYCH, The Lundbeck Foundation Initiative for Psychiatric Research, Copenhagen, DK 56, Brain and Mind Centre, University of Sydney, Sydney, NSW, AU

57, Interfaculty Institute for Genetics and Functional Genomics, Department of Functional Genomics, University Medicine and Ernst Moritz Arndt University Greifswald, Greifswald, Mecklenburg-Vorpommern, DE

58, Roche Pharmaceutical Research and Early Development, Pharmaceutical Sciences, Roche Innovation Center Basel, F. Hoffmann-La Roche Ltd, Basel, CH

59, Max Planck Institute of Psychiatry, Munich, DE

60, MRC Centre for Neuropsychiatric Genetics and Genomics, Cardiff University, Cardiff, GB

61, Department of Psychological Medicine, University of Worcester, Worcester, GB 62, Division of Research, Kaiser Permanente Northern California, Oakland, CA, US

63, Psychiatry & The Behavioral Sciences, University of Southern California, Los Angeles, CA, US

64, Department of Biomedical Informatics, Harvard Medical School, Boston, MA, US 65, Department of Medicine, Brigham and Women’s Hospital, Boston, MA, US

66, Informatics Program, Boston Children’s Hospital, Boston, MA, US

67, Wellcome Trust Centre for Human Genetics, University of Oxford, Oxford, GB

68, Institute of Social and Preventive Medicine (IUMSP), Lausanne University Hospital and University of Lausanne, Lausanne, VD, CH

69, Swiss Institute of Bioinformatics, Lausanne, VD, CH

70, Division of Psychiatry, Centre for Clinical Brain Sciences, University of Edinburgh, Edinburgh, GB

71, Mental Health, NHS 24, Glasgow, GB

72, Department of Psychiatry and Psychotherapy, University of Bonn, Bonn, DE 73, Statistics, University of Oxford, Oxford, GB

74, Psychiatry, Columbia University College of Physicians and Surgeons, New York, NY, US

75, School of Psychology and Counseling, Queensland University of Technology, Brisbane, QLD, AU

76, Child and Youth Mental Health Service, Children’s Health Queensland Hospital and Health Service, South Brisbane, QLD, AU

77, Child Health Research Centre, University of Queensland, Brisbane, QLD, AU 78, Estonian Genome Center, University of Tartu, Tartu, EE

79, Medical Genetics, University of British Columbia, Vancouver, BC, CA 80, Statistics, University of British Columbia, Vancouver, BC, CA

81, DZHK (German Centre for Cardiovascular Research), Partner Site Greifswald, University Medicine, University Medicine Greifswald, Greifswald, Mecklenburg-Vorpommern, DE

82, Institute of Clinical Chemistry and Laboratory Medicine, University Medicine Greifswald, Greifswald, Mecklenburg-Vorpommern, DE

83, Institute of Health and Biomedical Innovation, Queensland University of Technology, Brisbane, QLD, AU

84, Humus, Reykjavik, IS

85, Virginia Institute for Psychiatric & Behavioral Genetics, Virginia Commonwealth University, Richmond, VA, US

86, Clinical Genetics, Vrije Universiteit Medical Center, Amsterdam, NL 87, Complex Trait Genetics, Vrije Universiteit Amsterdam, Amsterdam, NL 88, Solid Biosciences, Boston, MA, US

89, Department of Psychiatry, Washington University in Saint Louis School of Medicine, Saint Louis, MO, US

90, Department of Biochemistry and Molecular Biology II, Institute of Neurosciences, Biomedical Research Center (CIBM), University of Granada, Granada, ES

91, Department of Psychiatry, University of Groningen, University Medical Center Groningen, Groningen, NL

92, Department of Psychiatry and Psychotherapy, University Hospital, Ludwig Maximilian University Munich, Munich, DE

93, Institute of Psychiatric Phenomics and Genomics (IPPG), University Hospital, Ludwig Maximilian University Munich, Munich, DE

94, Division of Cancer Epidemiology and Genetics, National Cancer Institute, Bethesda, MD, US

95, Behavioral Health Services, Kaiser Permanente Washington, Seattle, WA, US

96, Faculty of Medicine, Department of Psychiatry, University of Iceland, Reykjavik, IS 97, School of Medicine and Dentistry, James Cook University, Townsville, QLD, AU 98, Institute of Health and Wellbeing, University of Glasgow, Glasgow, GB

99, deCODE Genetics / Amgen, Reykjavik, IS

100, College of Biomedical and Life Sciences, Cardiff University, Cardiff, GB

101, Institute of Epidemiology and Social Medicine, University of Münster, Münster, Nordrhein-Westfalen, DE

102, Institute for Community Medicine, University Medicine Greifswald, Greifswald, Mecklenburg-Vorpommern, DE

103, Department of Psychiatry, University of California, San Diego, San Diego, CA, US 104, KG Jebsen Centre for Psychosis Research, Norway Division of Mental Health and Addiction, Oslo University Hospital, Oslo, NO

105, Medical Genetics Section, CGEM, IGMM, University of Edinburgh, Edinburgh, GB 106, Clinical Neurosciences, University of Cambridge, Cambridge, GB

107, Internal Medicine, Erasmus MC, Rotterdam, Zuid-Holland, NL

108, Roche Pharmaceutical Research and Early Development, Neuroscience, Ophthalmology and Rare Diseases Discovery & Translational Medicine Area, Roche Innovation Center Basel, F. Hoffmann-La Roche Ltd, Basel, CH

109, Department of Psychiatry and Psychotherapy, University Medicine Greifswald, Greifswald, Mecklenburg-Vorpommern, DE

110, Department of Psychiatry, Leiden University Medical Center, Leiden, NL

111, Virginia Institute for Psychiatric & Behavioral Genetics, Virginia Commonwealth University, Richmond, VA, US

112, Computational Sciences Center of Emphasis, Pfizer Global Research and Development, Cambridge, MA, US

113, Institute for Molecular Bioscience; Queensland Brain Institute, The University of Queensland, Brisbane, QLD, AU

114, Department of Psychiatry, University of Münster, Münster, Nordrhein-Westfalen, DE 115, Department of Psychiatry, Melbourne Medical School, University of Melbourne, Melbourne, AU

116, Florey Institute for Neuroscience and Mental Health, University of Melbourne, Melbourne, AU

117, Institute of Medical Genetics and Pathology, University Hospital Basel, University of Basel, Basel, CH

118, Institute of Neuroscience and Medicine (INM-1), Research Center Juelich, Juelich, DE 119, Amsterdam Public Health Institute, Vrije Universiteit Medical Center, Amsterdam, NL 120, Centre for Integrative Biology, Università degli Studi di Trento, Trento, Trentino-Alto Adige, IT

121, Department of Psychiatry and Psychotherapy, Medical Center - University of Freiburg, Faculty of Medicine, University of Freiburg, Freiburg, DE

122, Center for NeuroModulation, Faculty of Medicine, University of Freiburg, Freiburg, DE 123, Psychiatry, Kaiser Permanente Northern California, San Francisco, CA, US

124, Medical Research Council Human Genetics Unit, Institute of Genetics and Molecular Medicine, University of Edinburgh, Edinburgh, GB

125, Department of Psychiatry, University of Toronto, Toronto, ON, CA 126, Centre for Addiction and Mental Health, Toronto, ON, CA

127, Division of Psychiatry, University College London, London, GB

128, Neuroscience Therapeutic Area, Janssen Research and Development, LLC, Titusville, NJ, US

129, Institute of Molecular and Cell Biology, University of Tartu, Tartu, EE

130, Psychosis Research Unit, Aarhus University Hospital, Risskov, Aarhus, DK 131, Munich Cluster for Systems Neurology (SyNergy), Munich, DE

132, University of Liverpool, Liverpool, GB

133, Mental Health Center Copenhagen, Copenhagen Universtity Hospital, Copenhagen, DK 134, Human Genetics and Computational Biomedicine, Pfizer Global Research and Development, Groton, CT, US

135, Psychiatry, Harvard Medical School, Boston, MA, US 136, Psychiatry, University of Iowa, Iowa City, IA, US

137, Department of Psychiatry and Behavioral Sciences, Johns Hopkins University, Baltimore, MD, US

138, Department of Psychiatry and Psychotherapy, University Medical Center Göttingen, Goettingen, Niedersachsen, DE

139, Human Genetics Branch, NIMH Division of Intramural Research Programs, Bethesda, MD, US

140, Faculty of Medicine, University of Iceland, Reykjavik, IS

141, Child and Adolescent Psychiatry, Erasmus MC, Rotterdam, Zuid-Holland, NL 142, Psychiatry, Erasmus MC, Rotterdam, Zuid-Holland, NL

143, Psychiatry, Dalhousie University, Halifax, NS, CA

144, Division of Translational Epidemiology, New York State Psychiatric Institute, New York, NY, US

145, Department of Clinical Medicine, University of Copenhagen, Copenhagen, DK

146, Department of Medical & Molecular Genetics, King’s College London, London, GB 147, Psychiatry & Behavioral Sciences, Stanford University, Stanford, CA, US

148, NIHR Maudsley Biomedical Research Centre, King’s College London, London, GB 149, Genetics, University of North Carolina at Chapel Hill, Chapel Hill, NC, US

150, Psychiatry, University of North Carolina at Chapel Hill, Chapel Hill, NC, US

